# Association of Long-Term Air Pollution Exposure with Dementia-Related Neuropathologies at Autopsy in a Community-Based Cohort

**DOI:** 10.64898/2026.02.03.26345515

**Authors:** Tara E. Jenson, Ryan M. Andrews, Sara D. Adar, Lisa L. Barnes, David A. Bennett, Dustin Burnham, John Cursio, Amanda Gassett, Uschi Graham, Joel D. Kaufman, Melissa Lamar, David X. Marquez, Sukriti Nag, Günter Oberdörster, Marcia Pescador Jimenez, Julie A. Schneider, Adam A. Szpiro, Jayant M. Pinto, Jennifer Weuve

**Affiliations:** Boston University School of Public Health, Boston, MA, USA; University of Michigan School of Public Health, Ann Arbor, MI, USA; Department of Psychiatry and Behavioral Sciences, Rush University Medical Center, Chicago, Illinois, USA; Rush Alzheimer’s Disease Center, Rush University Medical Center, Chicago, Illinois, USA; Department of Environmental and Occupational Health Sciences, School of Public Health, University of Washington, Seattle, WA, USA; University of Illinois Chicago, Chicago, IL, USA; Pharmaceutical Sciences Department, College of Pharmacy, University of Kentucky, Lexington, KY 40536-0596, USA; Department of Pathology and Alzheimer’s Disease Center, Rush University Medical Center, Chicago, IL, USA; University of Rochester Medical Center, NY, USA; Department of Pathology (Neuropathology), Rush University Medical Center, Chicago, IL, USA; Department of Neurological Sciences, Rush University Medical Center, Chicago, IL, USA; University of Washington, Seattle, WA, USA; Section of Otolaryngology-Head and Neck Surgery, The University of Chicago, Chicago, IL, USA

## Abstract

**Objective:** To evaluate long-term antemortem exposure to four pollutants in relation to Alzheimer’s disease (AD), cerebrovascular, and other dementia-related neuropathologies, measured at autopsy.

**Design:** Retrospective cohort study.

**Setting:** Individual participant data from four Rush Alzheimer’s Disease Center (RADC) longitudinal cohort studies: Memory and Aging Project, Minority Aging Research Study, Rush Clinical Core, and Latino Core. The cohorts enrolled participants residing in Chicago, Illinois (USA), including its metropolitan area and suburbs, and in outlying areas of Illinois.

**Participants:** 909 decedents from the four RADC cohorts who underwent brain autopsy.

**Setting:** RADC cohorts are drawn from northeastern Illinois (IL) and Chicago metropolitan, suburban, and outlying areas of IL.

**Participants:** All participants were aged ≥60 years at enrollment. Analyses included 909 decedents with air pollution exposure measures who underwent autopsies prior to 2020 (of 3,579 who enrolled by the end of 2019); all autopsies were from community-based cohorts.

**Exposures:** Exposure to fine particulate matter (PM_2.5_; particles <2.5 μm in aerodynamic diameter), nitrogen dioxide (NO_2_), oxides of nitrogen (NO_x_; nitrous oxide and NO_2_ combined), and ground-level ozone (O_3_) during the five years preceding death. Exposures were estimated with validated models developed for both the conterminous USA and the Chicago metropolitan area.

**Main Outcomes and Measures:** Twelve dementia-related neuropathologies measured by a neuropathologist at autopsy: Alzheimer’s disease neuropathology (ADNC), β-amyloid density, tau tangle density, cerebral arteriolosclerosis, cerebral atherosclerosis, cerebral amyloid angiopathy, chronic cerebral infarctions (microscopic and gross), hippocampal sclerosis, Lewy bodies and limbic predominant age-related TDP-43 encephalopathy (LATE-NC).

**Results:** Exposure to PM_2.5_ and NO_2_, as measured using Chicago-specific models, were both associated with higher tau tangle density [mean difference per 2.5 µg/m^3^ PM_2.5_ = 0.25 tangles/mm^2^, (95% confidence interval [CI], -0.05 to 0.56); mean difference per 5 ppb NO_2_ = 0.10 tangles/mm^2^, (95% CI -0.07 to 0.28)]. PM_2.5_ exposure was associated with higher prevalence of arteriolosclerosis [prevalence ratio (PR) per 2.5 µg/m^3^ = 1.51 (95% CI, 1.02 to 2.24)]. Both PM_2.5_ and NO_x_ exposure were associated with higher prevalence of cerebral atherosclerosis [PR per 2.5 µg/m^3^ PM_2.5_ = 1.41 (95% CI, 0.93 to 2.13); PR per 5 ppb NO_x_ = 1.10 (95% CI, 0.98 to 1.23)]. None of the exposures was clearly adversely associated with the other neuropathologic outcomes, including β-amyloid density and ADNC.

**Conclusion and Relevance:** Higher exposure to PM_2.5_ was associated with cerebral arteriolosclerosis and atherosclerosis at death, consistent with the known vascular toxicity of this pollutant.

## INTRODUCTION

Long-term exposure to air pollution may contribute to the development of dementia.^1-3^ As such, air pollution is a modifiable exposure on a scale that could benefit entire populations through policy change. The dementia-related outcomes most studied with respect to exposure to air pollutants are cognitive function, dementia incidence, and cognitive decline, with the strongest evidence supporting an association of long-term fine particulate matter (PM_2.5_) exposure with cognitive decline. PM_2.5_ includes particles <2.5 μm in aerodynamic diameter as well as *ultrafine* particles (<0.1 μm) that, as demonstrated in animal models, appear to be able to translocate from the nasal passageto olfactory bulb and deeper brain regions.^4^ We recently reported evidence of this phenomenon in humans. In autopsied brains from the Rush Religious Orders Study, we identified numerous different metal, plastic and other particles, of likely exogenous origin, in olfactory bulb, amygdala, and cerebellum tissue.^5^ Evidence on nitrogen dioxide (NO_2_), oxides of nitrogen (NO_x_; nitrous oxide and NO_2_ combined), and ground-level ozone (O_3_) has been too inconsistent or sparse to be conclusive.^2^

The adverse effects of air pollution exposure in the periphery—cardiovascular and metabolic— are relevant to dementia etiology. ^6,7,8^ Air pollutants also may contribute to dementia etiology by inducing brain inflammation, oxidative damage, and microglial activation, as well as disturbing protein homeostasis.^6,7,8^ In particular, air pollutants have been hypothesized to promote specific neuropathologies. Much attention has focused on how air pollutants might promote Alzheimer’s disease (AD) neuropathology,^9-14^ i.e., the deposition of neurofibrillary tangles containing phosphorylated tau and neuritic plaques containing β-amyloid.^15-17^ However, other neuropathologies also contribute to dementia, e.g., Lewy body disease,^18^ hippocampal sclerosis,^19^ and limbic predominant age-related TDP-43 encephalopathy.^19,20^ They also include cerebrovascular pathologies.^21-25^ The potential effects of air pollution on cerebrovascular pathology are especially plausible, given the raft of evidence indicating that some air pollutants promote cardiovascular disease,^26-30^ including stroke.^30-32^

Indeed, dementias are often the result of combinations of multiple neuropathologies.^33-37^ In addition, among those with AD-related neuropathology, those who also have cerebrovascular pathologies are more likely to develop symptoms of dementia.^38,39^ Ultimately, understanding the role of air pollutants in promoting these non-AD neuropathologies can inform the effectiveness of exposure-reducing interventions, and, more generally, inform multi-pronged approaches to dementia prevention. There is a small body of evidence on air pollution exposure in relation to markers of dementia-related neuropathologies measured on autopsy.^3,40,41^ These studies have generated suggestive but inconsistent associations of antemortem exposure to PM_2.5_ with AD neuropathologies. There is considerably less evidence on other common air pollutants and other dementia-related neuropathologies. Therefore, for the Air Pollution and Alzheimer’s Dementia: Neuropathologic and Olfactory Mechanisms in Multi-Ethnic Longitudinal Cohorts (also known as AERONOSE) project, we linked residential history with up to 30 years of predicted air pollutant data, and evaluated the associations of long-term exposures to PM_2.5_, NO_2_, NO_x_ and O_3_ with Alzheimer’s disease (AD), cerebrovascular, and three other dementia-related neuropathology measures at autopsy among more than 900 participants from four cohorts of the Rush Alzheimer’s Disease Center.

## METHODS

### Study Sample

Data for this study was drawn from four Rush Alzheimer’s Disease Center (RADC) cohorts: the Rush Memory and Aging Project (MAP), Minority Aging Research Study (MARS), Rush Clinical Core (RCC), and Latino Core (LATC), all ongoing community-based cohorts of adults 60 years and older convened for the longitudinal study of dementia, and described in detail previously. ^42-45^ The cohorts collectively are drawn from northeastern Illinois (IL) and Chicago metropolitan, suburban, and outlying areas of IL. All participants undergo annual study-based clinical evaluation. Briefly, MAP began enrolling older adults without dementia from northeastern IL in 1997. ^42^ MARS began enrolling participants free of dementia in 2004 recruited from metropolitan Chicago and is composed primarily of African American participants.^43^ RCC, also composed primarily of African American participants, began enrollment in 2008 by recruiting patients, relatives and friends (irrespective of cognitive or dementia status) seen at the Rush Memory Clinic who presented for evaluation of cognitive concerns.^44^ LATC began in 2015 with enrollment of Latinx participants recruited in metropolitan Chicago and with no known dementia diagnosis.^45^ Agreement to autopsy and brain donation upon death is required for enrollment in MAP, but is voluntary for enrolling in the MARS, RCC and LATC cohorts.^42-45^ Our study included autopsy data from participants who were deceased by the end of 2019 to avoid including those who died from COVID-related causes. By the end of 2019, 3,579 total participants were enrolled across the MAP (N=2,208), MARS (N=791), RCC (N=344) and LATC (N=236) cohorts, with 987 total autopsies conducted (MAP N=885, with earliest autopsies in 1998; MARS N=33, with earliest autopsies in 2010; RCC N=68, with earliest autopsies in 2008; and LATC N=1, with earliest autopsies in 2017).^46^ Based on the availability of air pollution exposure measures, our analytical samples ranged from 749 to 909, as described below.

This study was approved by the Institutional Review Boards of Boston University and Rush University Medical Center.

## Exposure to air pollution

### Residential address history

Each participant’s exposure to air pollution was estimated based on their residential address locations during the time periods they lived at those locations along with the building blocks of air pollutant concentrations modeled over each serial two-week interval during their tenure at each location. Data on residential locations came from two sources: the RADC’s address records collected during annual in-home assessments for the duration of participation from enrollment through date of death, and LexisNexis. LexisNexis is a third-party commercial data services company that provides all known addresses for a requested set of individuals, using a proprietary algorithm that compiles address histories from sources that include financial transactions, credit records, Social Security Administration death records, voting records, vehicle licenses and registrations, and criminal records and legal filings.^47^ For this project, the “seed data” provided to the LexisNexis algorithm consisted of each participant’s first name, last name, and last known address. LexisNexis returned data containing up to 20 of the most recent addresses of each of 3,622 participants, the earliest and most recent dates associated with these addresses, and flags indicating if each address was a match to the seed data we provided, an updated address, or a non-match. The years covered spanned 1937-2023. We then cleaned and geocoded the LexisNexis address data using a combination of R and ESRI ArcGIS. RADC records covered participants’ known residential locations from study enrollment through date of death. The RADC reviewed all participant addresses to correct clerical errors in notations prior to applying internal geographic information systems (GIS) mapping via ESRI ArcGIS and US Census TigerLine data.^48^ We then concatenated the data from these two sources, favoring RADC address data for enrollment onward, with LexisNexis data used to fill in gaps and address history prior to enrollment. Each participant’s final resulting residential address history provided the spatial and temporal orientation for estimating their long-term exposure to air pollution.

### Exposure to PM_2.5_, NO_2_, NO_x_, and O_3_

We estimated each participant’s long-term exposure to fine particulate matter (PM_2.5_), nitrogen dioxide (NO_2_), oxides of nitrogen (NO_x_), and ground-level ozone (O_3_) by linking that participant’s address history to validated models that predict geographically specific air pollution concentrations on a fortnight basis, developed by the MESA Air Study.^31,49-53^ Chicago region-specific models predict air pollution concentrations of PM_2.5_, NO_2_, NO_x_ and O_3_ at a given location in the years from 1999-2018. Models for the contiguous United States predict concentrations of PM_2.5_ from 2000-2019, and for NO_2_ and O_3_ for 1990-2019. The Chicago-specific models are optimized for performance for the Chicago region, while the national pollutant prediction models incorporate all of the Chicago-specific monitor data, but effectively down-weight the Chicago data. As such measures computed from the national models have overall lower accuracy and precision for Chicago area locations than do measures computed from the corresponding Chicago-specific models. Given the differences in accuracy and precision in the pollutant measures modeled at the national vs Chicago levels, and the contrast in participant sample sizes available for national vs Chicago modeled pollutants, we include both national and Chicago-specific pollutant estimates in our study.

We then estimated each participant’s exposure to each air pollutant in the five years preceding their date of death, using the biweekly pollutant predictions linked with participant geocoded residential address history, and computed with the R intervalaverage package.^54^ We computed five-year exposures using each prediction model (referred to as Chicago or national). We adopted a five-year average pollutant exposure for our primary analyses in order to balance maximizing the length of the exposure term prior to death with maximizing our sample (Figure S1). Whether computing a given five-year exposure was possible depended on whether a participant’s residential location(s) and dates of residence fell within the model’s spatial and temporal coverage (Figure S1). Our final analytical sample sizes were N=909 for national model NO_2_ and O_3_ exposures, N=837 for national model PM_2.5_ exposure, N=775 for Chicago model NO_2_, NO_x_, and PM_2.5_ exposures, and N=749 for Chicago model O_3_ exposure (Figure S2). Note: differences in Chicago O_3_ sample size versus other Chicago exposures are due to differences in monitors detecting pollutant types.

### Assessment of dementia-related neuropathology

Preservation and evaluation procedures for autopsied brain tissues have been described previously.^42^ We evaluated 12 neuropathologic outcomes, using measures previously described in detail.

### Alzheimer’s disease-related

Level of Alzheimer’s disease pathology (ADNC) was determined by neuropathologist based on the National Institute on Aging-Alzheimer’s Association criteria that combine neurofibrillary tangles score (Braak), neuritic plaque score (CERAD), and beta-amyloid plaque score (Thal).^15^ ADNC was characterized as a four-level measure of AD likelihood: no AD, low likelihood, intermediate likelihood, or high likelihood. A dichotomized version of ADNC - AD present (intermediate or high likelihood; either level fulfills criteria for pathologic diagnosis of AD) or AD not present (no AD or low likelihood) - was used for this analysis.^15,46^ β-amyloid (Aβ) and tau tangle density were both quantified across 8 brain regions using procedures described previously,^16,46^ denoting a square root-transformed mean percent of Aβ positivity in the cortex and square root-transformed mean of tau-tangles per square mm, respectively.

### Cerebrovascular neuropathology

Cerebral atherosclerosis was measured using a four-level rating of large vessel cerebral atherosclerosis in seven areas of the brain.^21^ Presence of cerebral amyloid angiopathy in four neocortical regions was summarized as a semiquantitative measure.^23^ Arteriolosclerosis characterizes histological changes commonly found in the small vessels of the brain with older age.^22^ Cerebral atherosclerosis, cerebral amyloid angiopathy and arteriolosclerosis variables were each dichotomized for this analysis as none/mild or moderate/severe. Presence of a) one or more chronic gross cerebral infarctions, b) chronic microscopic infarctions, and c) one or more of any size chronic infarctions,^24,25^ and were dichotomously categorized for analysis as none or any.

### Other dementia-related pathologies

Presence of Lewy bodies was determined across seven brain regions^18^ and was dichotomized for analysis as present or absent. Presence of hippocampal sclerosis in the mid-hippocampus was determined^19^ and analyzed as present or absent. Presence of limbic predominant age-related TDP-43 encephalopathy (LATE-NC) inclusions in neurons and glia was determined across eight brain regions,^20^ and was analyzed dichotomously as: a) none or in amygdala only, or b) extending beyond the amygdala.

### Covariates

We adjusted pollutant exposure-neuropathology associations for several potential confounder and precision variables:^46^ post-mortem interval (interval between death and autopsy tissue preservation in hours); birth and death years to adjust for temporal trends in exposures; sex (self-reported as male, female); education (years); income at baseline and age 40 (<$30,000, $30,000-$49,999, $50,000-$74,999 and $75,000 or more); early-life socioeconomic status - a continuous composite index incorporating years of paternal and maternal education and number of children in family; apolipoprotein-ε4 carriership (no ε4 allele, 1-2 ε4 alleles); and race/ethnicity (Black, White, other race/ethnicity groups). Note: We did not further adjust for cohort, because the cohorts aligned strongly with the race/ethnicity groups we used in our analyses; that is, 99.9% of non-Latinx White participants were in MAP, and 72.6% of Latinx or Black participants were in the non-MAP cohorts. Given the low numbers of non-MAP participants, we imputed missing covariate data using median values of the overall sample.

### Statistical Analyses

#### Potential for selection bias

The data in our sample came from participants who passed through multiple stages of selection: consent to autopsy at enrollment (MAP, for which consent to autopsy is a requirement of enrollment) or enrollment followed by consent to autopsy (MARS, CC and LATC); death; and undergoing autopsy. These winnowing processes could have yielded an analytical sample in which the association of air pollution exposure with dementia-related neuropathology differed from that in the sample of RADC cohort members who could have been included had they died and undergone autopsy^55^ Prior to estimating the association of air pollution exposure with dementia-related neuropathology, we evaluated the potential for bias in our study from selection processes operating after enrollment. As detailed in **Appendix A: Evaluation of Potential for Selection Bias**, there was little empirical evidence to support the presence of selection bias from differential post-enrollment attrition, although selection bias from other processes remains possible.

#### Estimation of associations of air pollution exposure with dementia-related neuropathology

We estimated the associations of average exposure to each of the four air pollutants in the five years before death with each of the 12 dementia-related neuropathology outcomes. We generated estimates using pollutant exposures from both the national and Chicago-based models. We used linear regression models to estimate the mean difference per exposure increment in β-amyloid and tau tangle densities. We used log binomial regression models to estimate prevalence ratios per exposure increment of ADNC, cerebral arteriolosclerosis, cerebral atherosclerosis, cerebral amyloid angiopathy, vascular chronic infarcts (gross, microscopic, and total), Lewy bodies, hippocampal sclerosis, and LATE-NC, all dichotomized as described previously. All results were transformed to mean differences or prevalence ratios per 2.5 μg/m^3^ PM_2.5_, per 5 ppb NO_2_, per 5 ppb NO_x_, or per 2.5 ppb O_3_. These contrasts balance the range of air pollutant concentrations in our study with contrasts typically used in health effects studies of air pollution.

#### Sensitivity analyses

We conducted sensitivity analyses to assess the robustness of our primary findings. In these analyses, we estimated the associations of average air pollutant exposures on neuropathology outcomes: a) restricted to the subset of participants for whom we were able to estimate all seven national-modeled and Chicago-modeled five-year average exposures (N=730); and b) in which we used exposures estimated over the first three years of the five-year window prior to death, i.e., a two-year lag model using three-year average pollutant exposures two years prior to participant death (Figure S6).

All statistical analyses were performed in R Studio (version 2025.09.2+418).

## Results

### Descriptive statistics

In the five years preceding their deaths, the median (IQR) exposure to PM_2.5_ among the cohort decedents was 9.9 (8.9-11.4) μg/m^3^, as predicted by national models (N=837; Table 1). The analogous medians (IQRs) were 11.0 (8.4-13.8) ppb for NO_2_ (N=909) and 23.0 (21.6-24.5) ppb for O_3_ (N=909).

**Table 1.**
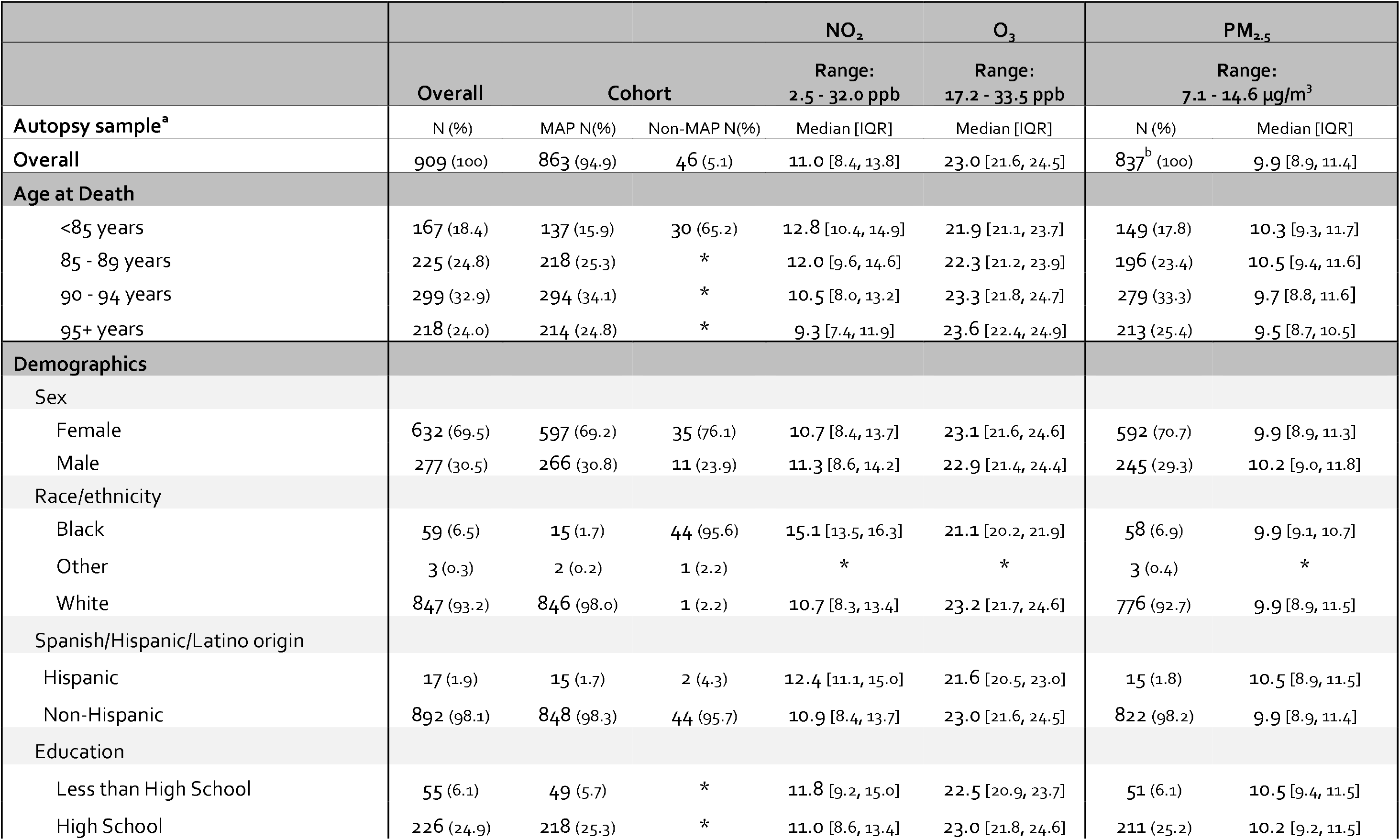
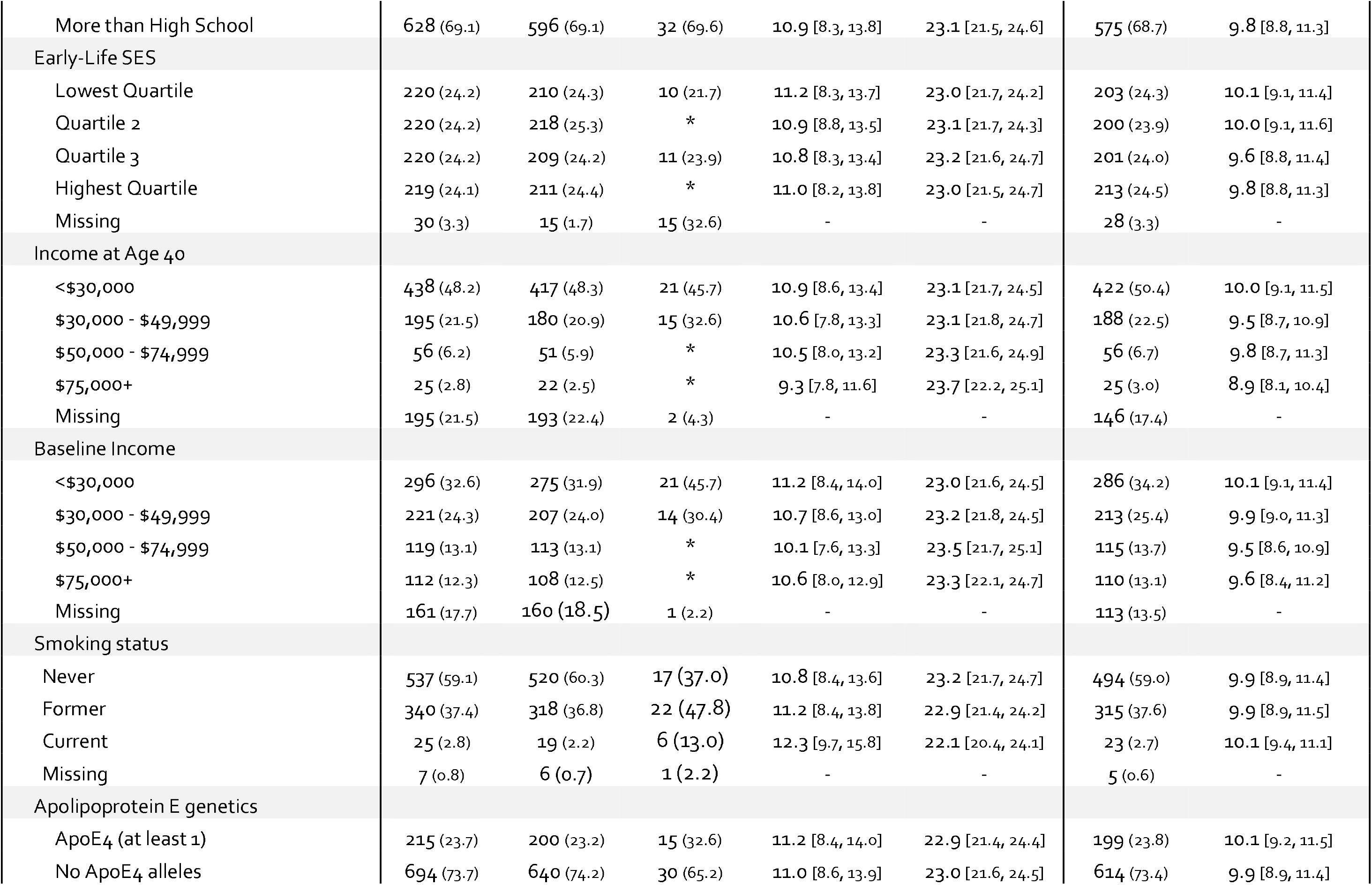
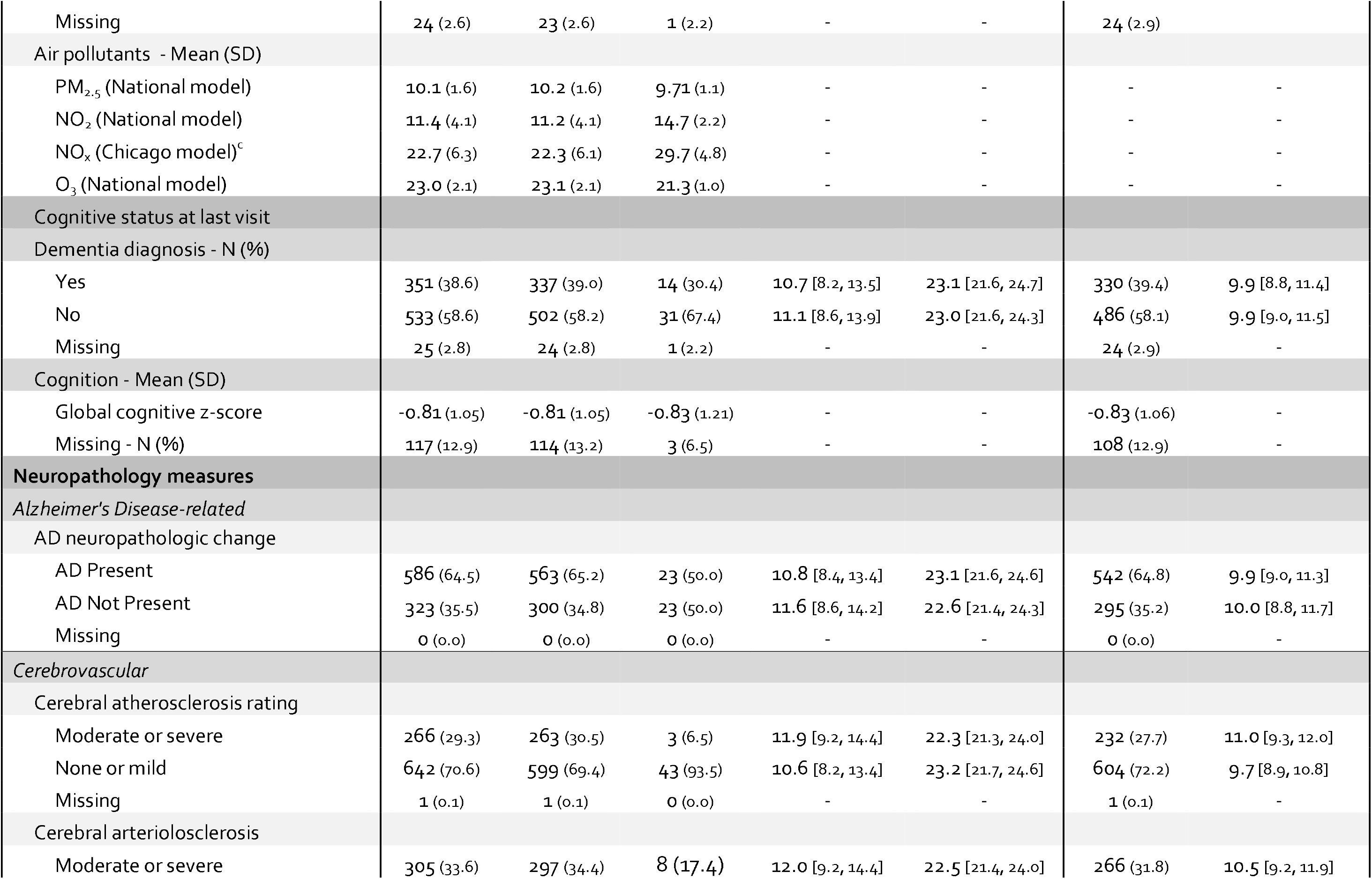
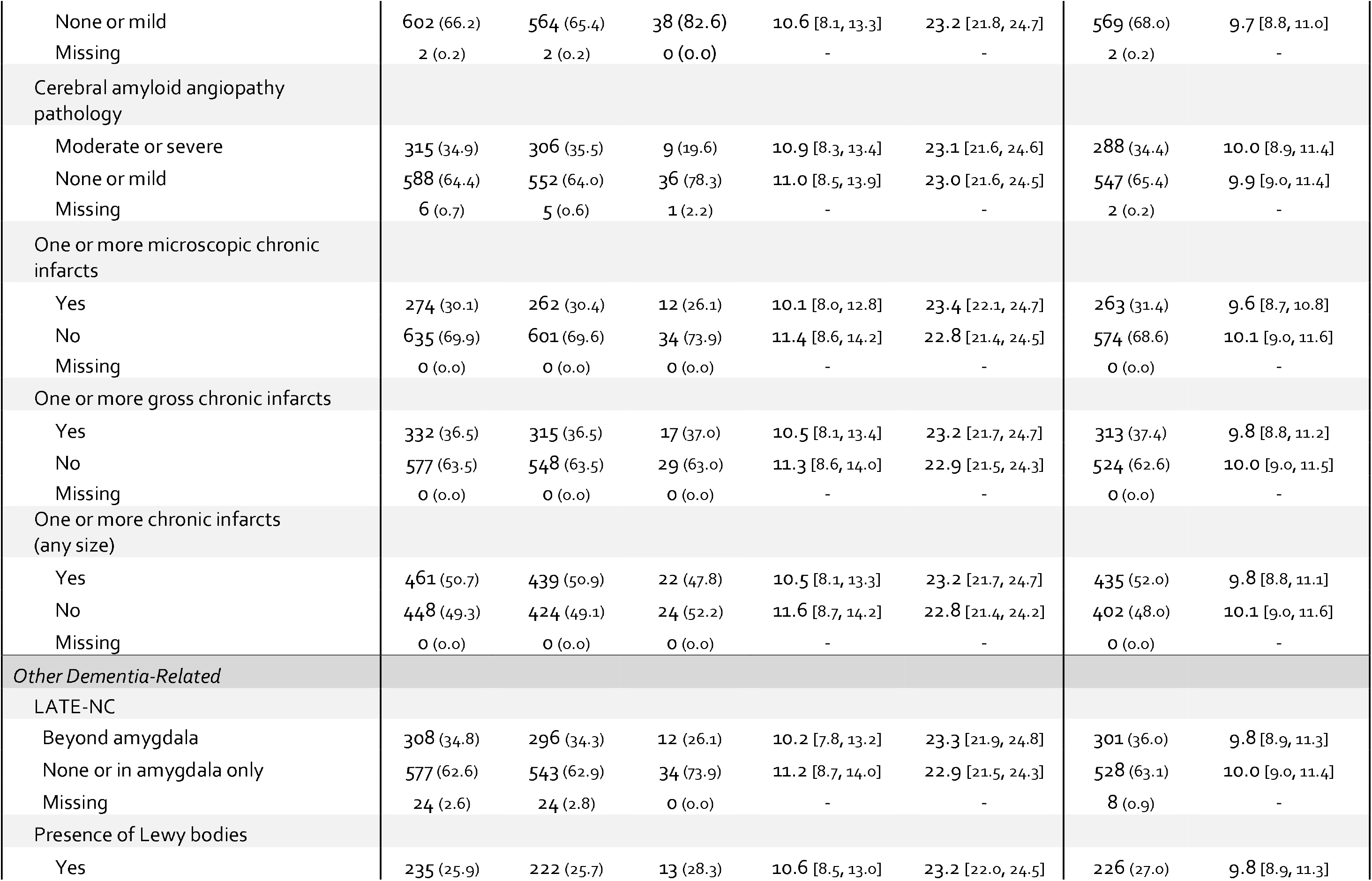
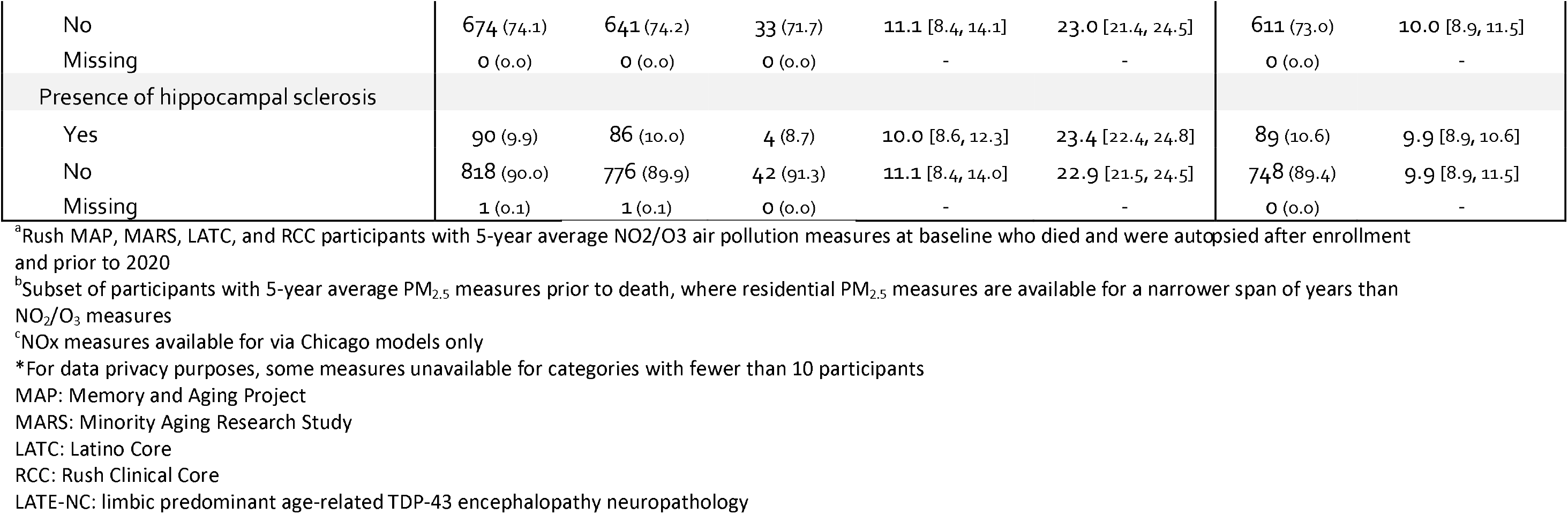
Median and IQR five-year average exposures to PM_2.5_, NO_2_, NO_x_, and O_3_* prior to death, by characteristics of the combined Rush MAP, MARS, RCC, and LATC autopsy cohort.

Five-year average antemortem exposure to PM_2.5_, NO_2_, and NO_x_ declined over time (Figures S5 and S6), tracking with known secular trends.^56,57^ Chicago-modeled PM_2.5_ and NO_2_ exposures slightly exceeded their nationally modeled counterparts (Figures S7, S8). By contrast, five-year average antemortem O_3_ generally increased over time (Figure S9), not unexpected given the countervailing trends in PM_2.5_ and NO_2_ and the atmospheric chemistry that generates O_3_ as a secondary pollutant (Figure 1).^58-60^

**Figure 1.**
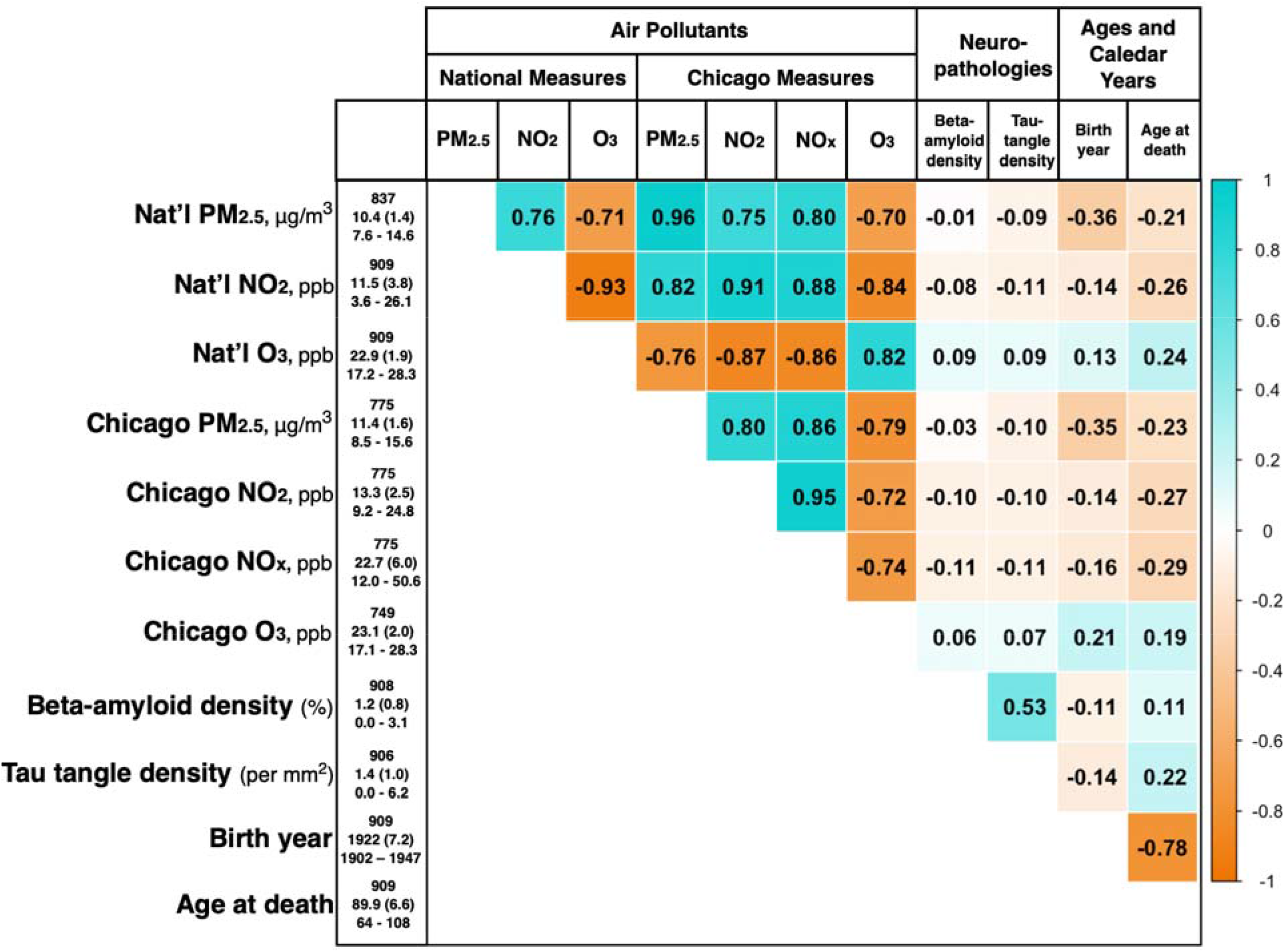
Distributions of and Correlations between estimated Five-Year Average Exposure to Select Air Pollutants, Neuropathologies, and Temporal Variables. RADC Cohorts: MAP, MARS, LATC, RCC. RADC: Rush Alzheimer’s Disease Center. MAP: Memory and Aging Project. MARS: Minority Aging Research Study. LATC: Latino Core, RCC: Rush Clinical Core.

Of the 10 binary neuropathologic outcomes we examined, the most common was ADNC (65% with Alzheimer’s indicated as present; Table 1). Cerebrovascular pathology was common as well, with about half of decedents having one or more chronic infarcts, and moderate to severe cerebrovascular pathology present in 28-35%. More than one-third of decedents had LATE-NC pathology extending beyond the amygdala. Less prevalent were Lewy bodies (26-27%) and hippocampal sclerosis (10-11%).

In unadjusted comparisons of median and IQRs, 5-year pollutant exposures were not strongly associated with many key characteristics of the participants (Table 1). For example, median 5-year concentrations of nationally modeled NO_2_, O_3_, and PM_2.5_ did not differ markedly by antemortem prevalence of dementia (Table 1). By contrast, older age at death corresponded to lower NO_2_ and PM_2.5_, but higher O_3_ exposure (Table 1, Figure 1). Compared with White decedents, Black decedents had higher antemortem NO_2_ exposure and lower antemortem O_3_ exposure. Antemortem exposure to O_3_ was positively correlated with educational attainment.

### Associations of air pollutant exposure with dementia-related neuropathology

Air pollutant exposure in the five years preceding death was generally not associated with higher postmortem burden of Alzheimer’s dementia-related neuropathology in multivariable-adjusted analyses (Figure 2; Tables S3 and S4). A few specific associations of air pollutant exposure were consistent with higher pathologic amyloid and, separately, pathologic tau. For example, higher Chicago-modeled O_3_ corresponded to higher β-amyloid density (mean difference per 2.5 ppb=0.07%, 95% CI -0.02, 0.15), while higher PM_2.5_ and NO_2_ were both consistent with higher tau tangle density (mean difference per 2.5 µg/m^3^ Chicago-modeled PM_2.5_=0.25/mm^2^, 95% CI -0.05, 0.56; mean difference per 5 ppb NO_2_=0.10/mm^2^, 95% CI -0.07, 0.28)). Puzzlingly, exposure to PM_2.5_ was strongly and inversely associated with β-amyloid density. The analogous associations involving exposures predicted by national models were effectively null. Associations of pollutants with ADNC, a measure combining features of pathologic amyloid and tau neuropathology, were small and estimated imprecisely (Figure 2, Table S4).

**Figure 2.**
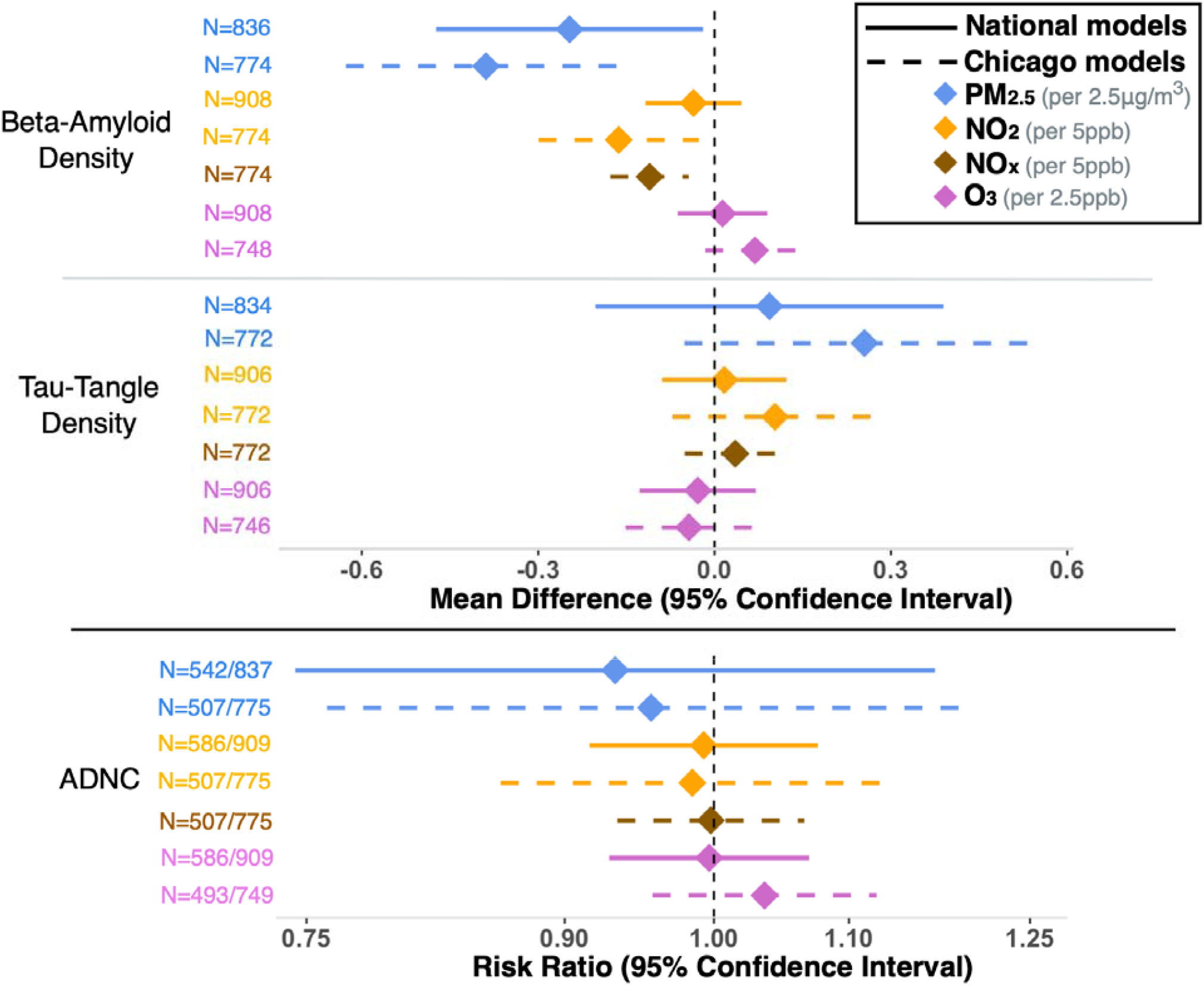
Association between 5-year average exposure to national- and Chicago-modeled air pollutants and AD-related neuropathologies at autopsy. RADC Cohorts: MAP, MARS, LATC, RCC. Exposures computed over the 5 years prior to death, using national and Chicago prediction models. N = events/total sample size for dichotomous outcomes or sample size for continuous outcomes. Beta-amyloid (Aβ) density was measured as the square root-transformed mean percent of Aβ positivity in the cortex. Tau tangle density was measured as square root-transformed mean of tau-tangles per square mm. We used linear regression models to estimate the mean difference per exposure increment in β-amyloid and tau tangle densities. Models were adjusted for time to autopsy, birth year, death year, sex, race/ethnicity, years of education, baseline smokin status, baseline income, income at age 40, early life socioeconomic status, ≥ 1 ApoE4 allele. ADNC: Alzheimer’s disease neuropathology. RADC: Rush Alzheimer’s Disease Center. MAP: Memory and Aging Project. MARS: Minority Aging Research Study. LATC: Latino Core, RCC: Rush Clinical Core.

Antemortem exposure to air pollution was more consistently associated with postmortem cerebrovascular pathology, particularly cerebral arteriolosclerosis (Figure 3; Table S4). The association involving PM_2.5_ exposure was most pronounced; for example, a 2.5-µg/m^3^ increment in PM_2.5_ exposure via the Chicago-based model corresponded to 51% greater prevalence in arteriolosclerosis (95% CI, 1.02, 2.24). Smaller positive associations were also present for NO_x_ and NO_2_; for example, the arteriolosclerosis prevalence ratio (PR) per 5-ppb increment in 5-year NO_x_ exposure was 1.09 (95% CI, 0.98, 1.22). Counter to expectation, O_3_ exposure was inversely associated with this outcome. Exposures to PM_2.5_, at least as measured by the Chicago-based model, and to NO_x_ were also associated with higher prevalence of cerebral atherosclerosis (PR per 2.5 µg/m^3^ PM_2.5_ = 1.41 [95% CI, 0.93, 2.13]; PR per 5 ppb NO_x_ = 1.10 [95% CI, 0.98, 1.23]). Although higher exposure to O_3_ was associated with the presence of one or more gross chronic infarctions (PR per 2.5 ppb O_3_ via the Chicago model=1.11 [(95% CI, 0.97, 1.28]; Figure 3, Table S5), this exposure was not associated with chronic microscopic infarctions. Unexpectedly, higher PM_2.5_ exposure was suggestively but inversely associated with gross chronic infarctions (Figure 3, Table S5). None of the air pollutant exposures were markedly adversely associated with cerebral amyloid angiopathy.

**Figure 3.**
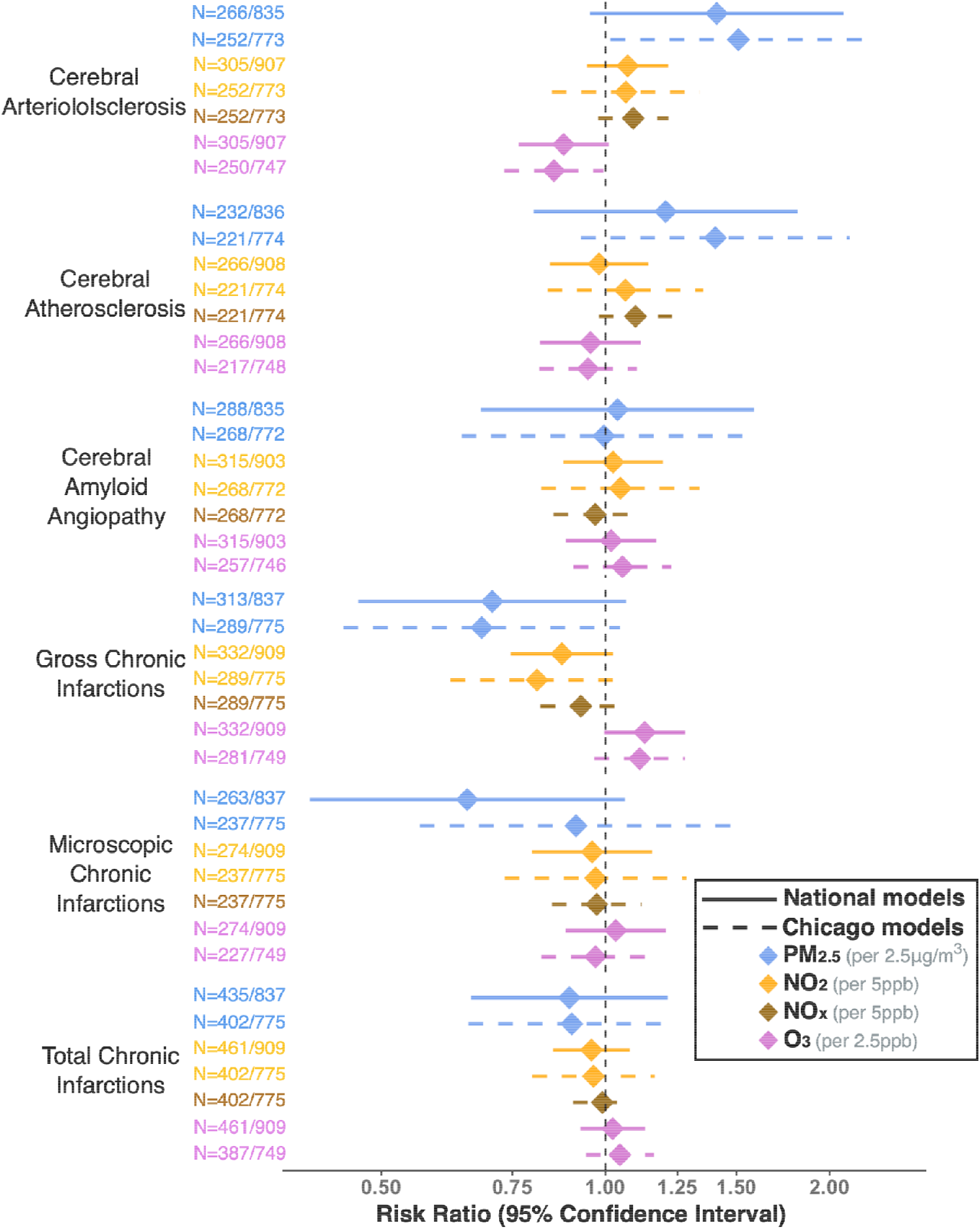
Association between 5-year average exposure to national- and Chicago-modeled air pollutants and cerebrovascular neuropathologies at autopsy. RADC Cohorts: MAP, MARS, LATC, RCC. Exposures computed over the 5 years prior to death, using national and Chicago prediction models. N = events/total sample size for dichotomous outcomes. Models were adjusted for time to autopsy, birth year, death year, sex, race/ethnicity, years of education, baseline smoking status, baseline income, income at age 40, early life socioeconomic status, ≥ 1 ApoE4 allele. AD: Alzheimer’s disease. RADC: Rush Alzheimer’s Disease Center. MAP: Memory and Aging Project. MARS: Minority Aging Research Study. LATC: Latino Core, RCC: Rush Clinical Core

There were no clear positive associations of any air pollutant exposure with postmortem prevalence of Lewy body disease, LATE-NC, or hippocampal sclerosis (Figure 4, Table S6).

**Figure 4.**
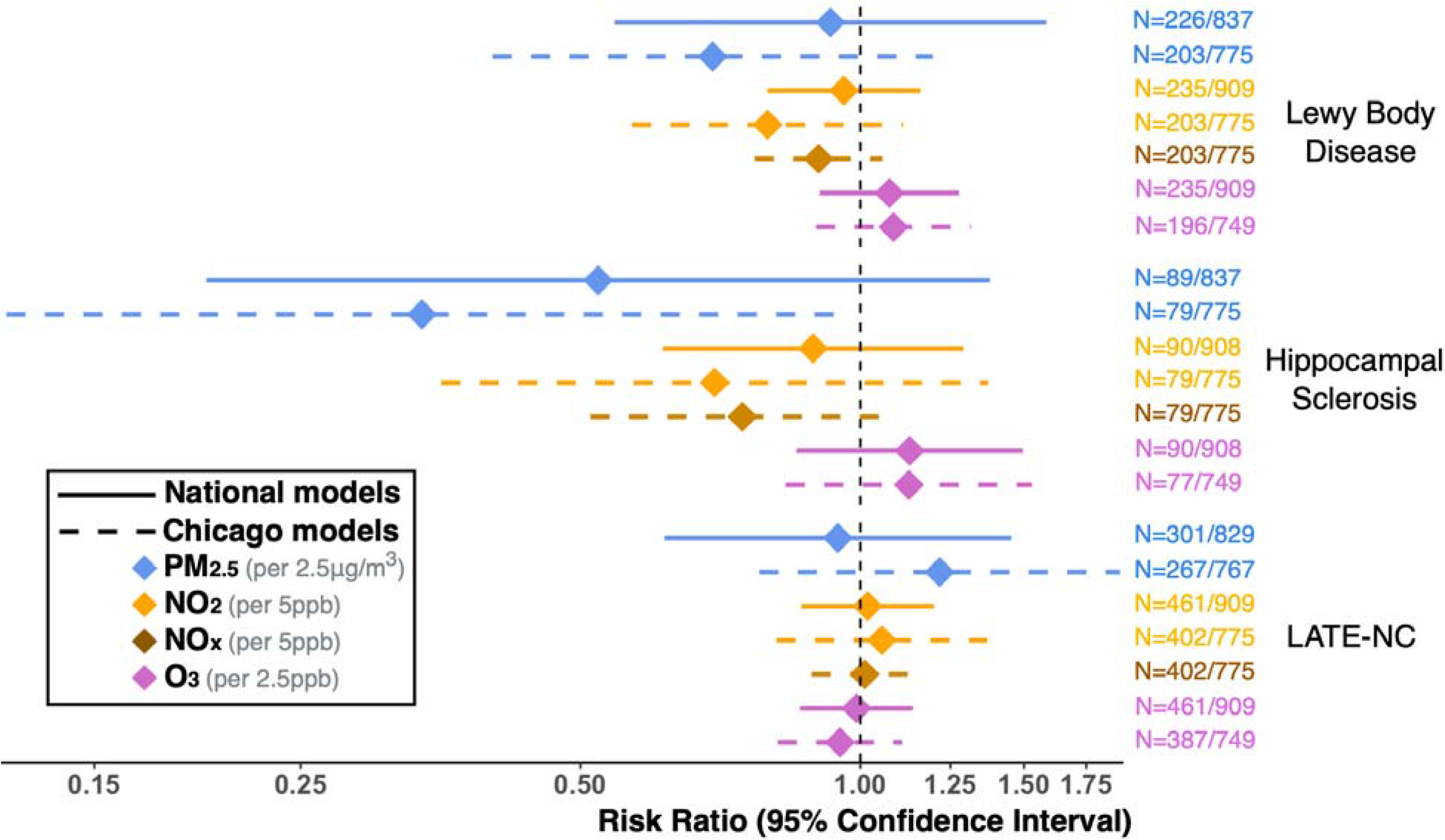
Association between 5-year average exposure to national and Chicago modeled air pollutants and other dementia-related neuropathologies at autopsy. RADC Cohorts: MAP, MARS, LATC, RCC. Exposures computed over the 5 years prior to death, using national and Chicago prediction models. N = events/total sample size for dichotomous outcomes. Models were adjusted for time to autopsy, birth year, death year, sex, race/ethnicity, years of education, baseline smoking status, baseline income, income at age 40, early life socioeconomic status, ≥ 1 ApoE4 allele. AD: Alzheimer’s disease. RADC: Rush Alzheimer’s Disease Center. MAP: Memory and Aging Project. MARS: Minority Aging Research Study. LATC: Latino Core, RCC: Rush Clinical Core; LATE-NC: limbic predominant age-related TDP-43 encephalopathy neuropathology.

The sensitivity analyses in which we a) restricted to the subset of participants for whom we were able to estimate all seven national-modeled and Chicago-modeled five-year average exposures (Tables S7 and S8); and b) used exposures estimated over the first three years of the five-year window prior to death (Tables S9 and S10) generated findings consistent with results from our primary analyses.

## DISCUSSION

We evaluated long-term exposure to four federally regulated air pollutants—PM_2.5_, NO_2_, NO_x_ and O_3_—in relation to AD-related, cerebrovascular and other dementia-related neuropathologies at autopsy in a large, well-characterized sample. Among 909 autopsied participants drawn from four cohort studies, higher exposure to PM_2.5_ was associated with substantially elevated prevalence of cerebral arteriolosclerosis. For example, a 2.5-µg/m^3^ increment in exposure corresponded to a 51% higher prevalence when we estimated exposure using a spatiotemporal prediction model built specifically for the Chicago metropolitan area. Higher NO_x_ and NO_2_ exposures were also adversely associated with this outcome, though to a lesser degree. We also observed adverse associations of exposures to PM_2.5_ and NO_x_ with cerebral atherosclerosis. Higher O_3_ exposure was associated with the presence of gross chronic infarctions. By and large, we found little evidence clearly indicating adverse associations of any of the pollutant exposures with neuropathologic Alzheimer’s disease. Associations with composite measures of AD neuropathologic burden and associations with measures of beta-amyloid and pathologic tau were null or inconsistent depending on whether we estimated exposure with national or Chicago-specific models.

AERONOSE marks one of the first investigations of air pollution exposure in relation to a wide-ranging set of dementia-related neuropathologies on autopsy, and one of the first such autopsy studies to examine antemortem exposure to NO_2_, NO_x_, O_3_, in addition to PM_2.5_, a pollutant that has been evaluated in three previous autopsy studies.^3,40,41^ Our findings on antemortem PM_2.5_ exposure and composite measures of AD-related neuropathology add to mixed findings from these three prior studies, which reported on the ABC score (combined measure of neuritic plaques and neurofibrillary tangles),^3,40^ and pathological diagnosis of AD measures that combined measures of neuritic plaques, diffuse plaques, and neurofibrillary tangles.^15,17^ Similar to our study, Shaffer et al. found no association of 10-year antemortem exposure to PM_2.5_ with ABC score, using data from the Adult Changes in Thought Study (ACT), a community-based cohort, and a sample size comparable to ours.^3^ Two smaller studies reported positive associations, by contrast. One study entailed 602 autopsies from a clinical cohort, composed entirely of individuals clinically diagnosed with AD or Parkinson’s disease, based at the Center for Neurodegenerative Disease Research at University of Pennsylvania.^41^ In this study, Kim et al. found a positive association of one-year antemortem exposure to PM_2.5_ with ADNC (odds ratio [OR] per 1 μg/m^3^, 1.19; 95% CI, 1.11 to 1.28).^40^ Similarly, Christensen et al. reported a positive association of one-year antemortem PM_2.5_ exposure with ABC score, based on data from a brain tissue donated by 224 decedents from community-based and clinical cohorts based at the Emory Goizueta AD Research Center.^40^

In all three previous studies, higher antemortem exposure to PM_2.5_ was associated with density of neuritic plaques. For example, in the ACT autopsy data, each 1-μg/m^3^ increment in 10-year PM_2.5_ exposure corresponded to a 35% higher odds of worse neuritic plaque density (e.g. none vs sparse, moderate or frequent) measured by Consortium to Establish a Registry for Alzheimer’s disease (CERAD) neuropathology score (OR per 1 μg/m^3^, 1.35, 95% CI 0.90, 1.90).^3^ In the Emory data, each 1-µg/m^3^ increment in one-year average PM_2.5_ exposure preceding death was associated with nearly twice the odds of worse CERAD score (OR 1.92; 95% CI 1.12– 3.30).^40^ The University of Pennsylvania reported an OR for worse CERAD score data of 1.20 (95% CI, 1.11 to 1.30), and an OR of 1.17 (95% CI 1.08, 1.27) for higher (worse) Thal amyloid phase per 1-µg/m^3^ increment in one-year PM_2.5_ exposure.^41^ Results from these studies contrast with our finding of no positive association between five-year exposure to PM2.5 and β-amyloid density, a measure comparable to CERAD score and Thal amyloid phase neuropathologies.

The suggestion in AERONOSE that higher PM_2.5_ exposure (Chicago model) is associated with higher tau tangle density corresponds with findings from the Emory brain bank, which also reported associations, albeit imprecise, of higher antemortem PM_2.5_ exposure with more advanced Braak stage (a neuropathology measure comparable to tau tangle density),^40^ and from the University of Pennsylvania data in which higher one-year average antemortem PM_2.5_ exposure corresponded to worse Braak stage (OR per 1-µg/m^3^=1.20; 95% CI 1.11, 1.29).^41^ In ACT, however, antemortem PM_2.5_ exposure was not associated with Braak stage.^3^

The strong positive association of PM_2.5_ exposure with arteriolosclerosis in AERONOSE is consistent with PM_2.5_ contributing to cerebral small vessel disease. Small vessel disease appears to promote tau pathology through pathways independent from beta-amyloid deposition.^61,62^ With pathologic tau as a downstream effect of arteriolosclerosis, this could explain the weak positive association we observed of PM_2.5_ exposure with tau pathology in the absence of a positive association with beta-amyloid. The University of Pennsylvania study, by contrast, observed no association between PM_2.5_ exposure and moderate to severe arteriolosclerosis (OR per 1-µg/m^3^ increment in PM_2.5_, 1.01; 95% CI, 0.92, 1.10).^41^ Of note, this study measured arteriolosclerosis in the occipital white matter of individuals diagnosed with clinical AD or Parkinson’s disease, with about 16% of decedents showing this pathology. In our study, arteriolosclerosis was measured in the anterior basal ganglia irrespective of antemortem diagnosis, and the prevalence was 33%. These distinctions could speak to the selection of the participants, or to the typical pattern of arteriolosclerosis spread through the brain, which has been hypothesized (though not confirmed) to start in the basal ganglia.^63^

In AERONOSE, exposure to PM_2.5_ was not associated with chronic gross or chronic microscopic infarctions, whereas it was positively associated with large infarctions (upon gross and microscopic examination) in the University of Pennsylvania study (OR per 1-µg/m^3^ PM_2.5_, 1.16; 95% CI, 1.01, 1.32).^41^ Neither study observed associations of PM_2.5_ exposure with cerebral amyloid angiopathy, Lewy body disease and LATE-NC stage.^40,41^

Estimating the relation of air pollution exposure to dementia-related neuropathology in humans is challenging. Although some neuropathologies can now be measured with antemortem imaging and biomarkers in blood and cerebral spinal fluid, brain autopsy still provides the most expansive slate of neuropathologies and their locations. Yet autopsy datasets are scarce and are usually small. Sample sizes ranged from 224 to 909 in the now four studies, including ours, of air pollution and autopsy-based neuropathology.

Moreover, processes of selection may yield study samples that do not represent the true relation of air pollution to neuropathology in the general population of older adults. For example, the University of Pennsylvania sample included only individuals clinically diagnosed with AD or Parkinson’s disease. The Emory sample also drew from an indeterminate number of clinical sources. The AERONOSE and ACT samples, by contrast, were drawn solely from community-based cohort studies.

Autopsy in these studies required both antemortem consent and death. If these steps are jointly related to air pollution exposure and neuropathology, there is potential for selection bias. There are known sociocultural differences in willingness to consent to brain autopsy at death.^64-66^ About 93% of our sample identified as White, in contrast with 75% of U.S. adults older than 65 years of age identifying as non-Hispanic White.^67^ In both ACT and AERONOSE, the combined endpoint of death and autopsy was more common among people who developed dementia.^68^ Nonetheless, both decedent samples represent unusually healthy segments of the population, with mean ages at death of 89 (ACT) and 91 years (AERONOSE). Finally, in the general population, both dementia and exposure to the pollutants we studied are associated with higher risk of death,^26-28,69,70^ the event that necessarily precedes autopsy. In the ACT study, Shaffer et al. accounted for potential bias emanating from differential selection between enrollment through autopsy by deploying inverse probably-of-continuation weights.^3^ Using a principled approach, we found little evidence to suggest differential selection between enrollment and autopsy (Appendix A). Nonetheless, in our study (as described in Appendix A) as in all of these studies, there remains the possibility of differential selection into the cohorts from which the autopsy samples were drawn.^71,72^

A second challenge of air pollution-brain autopsy studies concerns measuring exposure to air pollution. We measured participants’ long-term exposure to air pollution using spatiotemporal prediction models that were developed for the conterminous United States and for the Chicago metropolitan area. The national models allowed us to include data from participants who resided outside of the Chicago area for some or all of the five-year period preceding their deaths. Nonetheless, most participants (N=775) lived in the Chicago area, and the highly spatially and temporally resolved Chicago prediction model was well-suited for distinguishing exposures in that area. This resolution may have conferred more accuracy to the exposures estimated from the Chicago model, possibly explaining why positive associations of pollutant exposures with autopsy outcomes were larger when we estimated exposures with the Chicago model. Most of the participants in our study had PM_2.5_ exposure that exceeded 9 µg/m^3^, the current U.S. federal regulatory standard for annual concentration.^73^ In addition, the ranges of exposure to all four pollutants were wide, enhancing our ability to observe associations. The ACT and University of Pennsylvania studies had ranges of PM_2.5_ exposure comparable to ours. One-year PM_2.5_ exposure in the Emory study was much lower (median, 1.32 µg/m^3^) with less variation (interquartile range of 0.18–3.77 µg/m^3^).

The timing and duration of exposure that is relevant for dementia etiology is unknown. Dementia is a late, symptomatic stage of pathophysiologic processes that have developed progressively over decades.^74^ Air pollution likely contributes to several steps in that process, and long-term exposure would capture most of those contributions. Estimating long-term exposure using spatiotemporal prediction models requires that those models cover long periods—ideally decades—along with participants’ address histories. The ACT study measured PM_2.5_ exposure over the 10 years preceding death and found similar results in sensitivity analyses of 20-year exposure. The five-year exposures in our study, by contrast, are unlikely to reflect effects on the early stages of neuropathology development, except to the extent that these exposures serve as a surrogate for longer-term exposure.

In this study of long-term exposure to four air pollutants in relation to 12 measures of dementia-related neuropathology, higher exposure to PM_2.5_ was strongly associated with risk of cerebral arteriolosclerosis, consistent with the known vascular toxicity of PM_2.5_. In contrast, none of the pollutants was consistently associated with higher risk of neuropathologic AD, LATE-NC, Lewy bodies, or hippocampal sclerosis.

## Data Availability

All dataset used in this study are publicly available by request from Rush Alzheimer's Disease Center (RADC) Research Resource Sharing Hub.

https://www.radc.rush.edu/

