## Supplementary Materials for "Association of Long-Term Air Pollution Exposure with Dementia-Related Neuropathologies at Autopsy in a Community-Based Cohort"

**Table S7**. Results from sensitivity analysis (a). Estimated mean difference and 95% confidence intervals (CI) in beta-amyloid density and tau tangle density at autopsy per increment in 5-year average exposure to national- and Chicago-modeled air pollutants, restricted to the subset of 730 participants for whom we were able to estimate all seven national-modeled and Chicago-modeled five-year average exposures ............................................................................... page 29

**Table S8.** Results from sensitivity analysis (a). Risk ratios (RR) and 95% confidence intervals (CI) for association of 5-year average exposure to national- and Chicago-modeled air pollutants with ADNC, cerebral arteriolosclerosis, cerebral atherosclerosis, cerebral amyloid angiography, chronic cerebrovascular infarctions, Lewy bodies, hippocampal sclerosis, and LATE-NC at autopsy, restricted to the subset of 730 participants for whom we were able to estimate all seven national-modeled and Chicago-modeled five-year average exposures ................... page 30

**Table S9.** Results from sensitivity analysis (b). Estimated mean difference and 95% confidence intervals (CI) in beta-amyloid density and tau tangle density at autopsy per increment in 5-year average exposure to national- and Chicago-modeled air pollutants, in which we used exposures estimated over the first three years of the five-year window prior to death .................... page 31

**Table S10.** Risk ratios (RR) and 95% confidence intervals (CI) for association of 5-year average exposure to national- and Chicago-modeled air pollutants with ADNC, cerebral arteriolosclerosis, cerebral atherosclerosis, cerebral amyloid angiography, chronic cerebrovascular infarctions, Lewy bodies, hippocampal sclerosis, and LATE-NC at autopsy, in which we used exposures estimated over the first three years of the five-year window prior to death .................................................................................................................................. page 33

**Figure S1. Construction of 5-year air pollution exposure windows prior to participant death for select air pollutants (RADC cohorts MAP, MARS, LATC, RCC).**

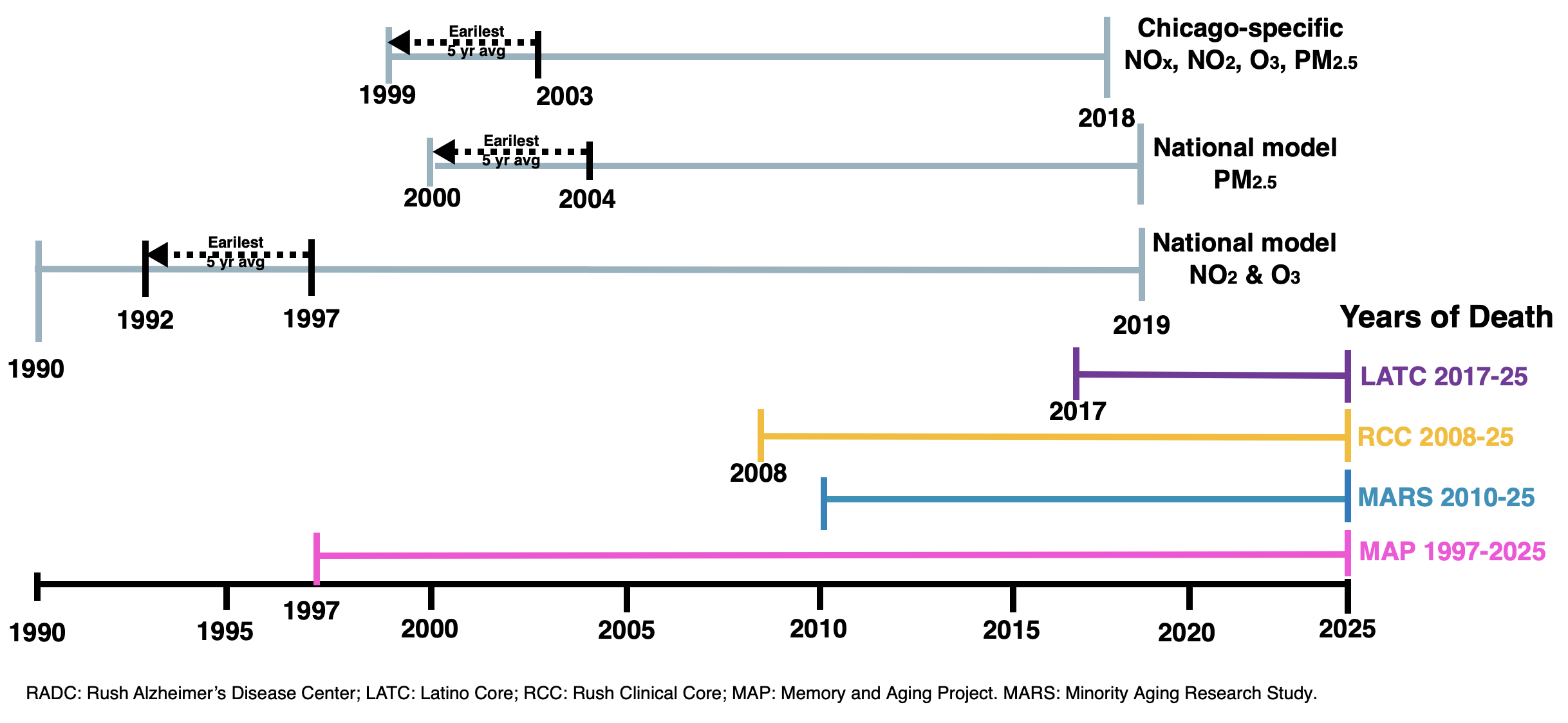

**Figure S2. Inclusion for air pollution and neuropathology models for select air pollutants, RADC Cohorts (MAP, MARS, LATC, RCC cohorts).**

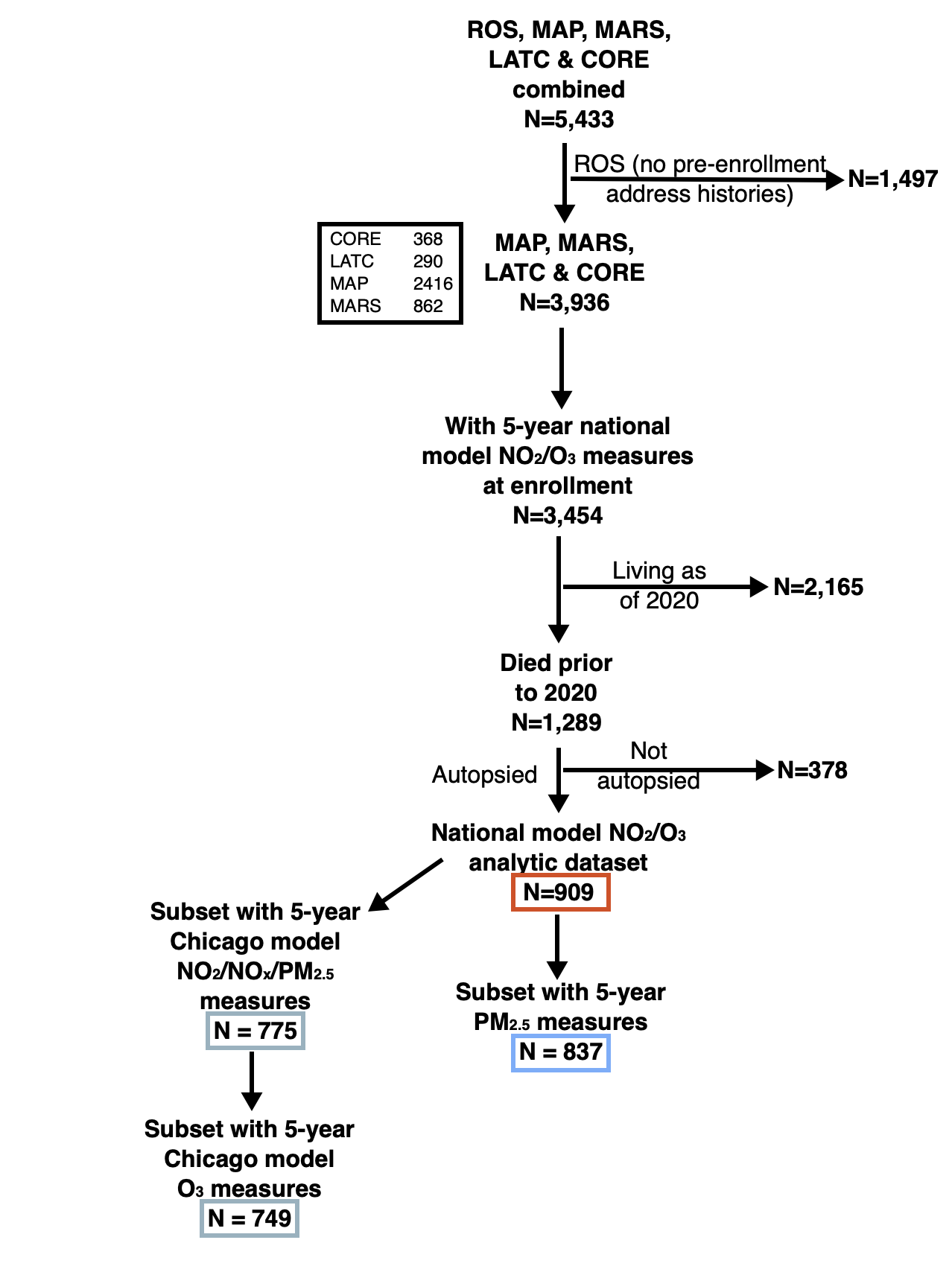

RADC: Rush Alzheimer’s Disease Center; LATC: Latino Core; CORE/RCC: Rush Clinical Core; MAP: Memory and Aging Project. MARS: Minority Aging Research Study.

**Appendix A: Evaluation of Potential for Selection Bias**

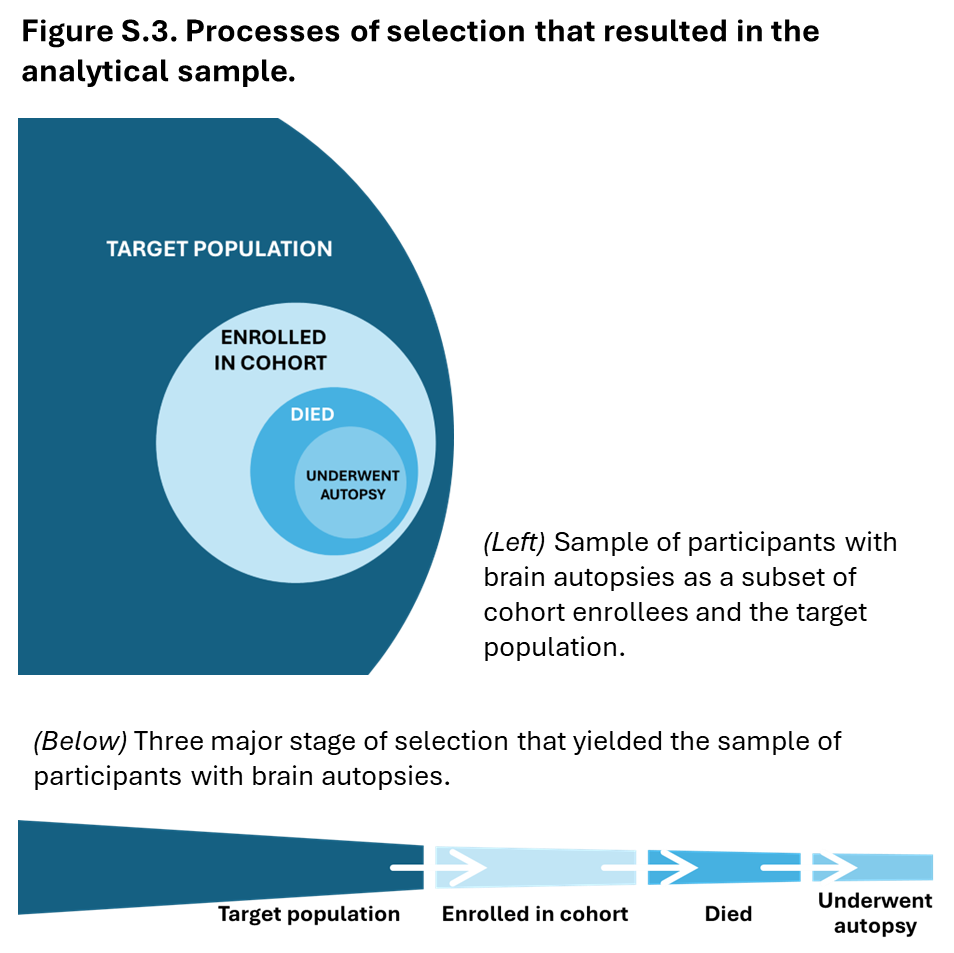
When the effect of an exposure estimated in a sample differs from the true effect in its source population, for reasons solely due to who was included (i.e., not confounding, measurement error, immortal time, or sampling error), we call this selection bias.^1^ It is possible that selection bias influenced the effects estimated in our study. The participants whose data were in our primary analyses were a sample of the target population of older adults living in the United States in the 1990 through 2019 who were at risk for developing dementia-related neuropathologies. Specifically, those participants passed through three major stages of selection for their data to be included: from the target population to enrolling in one of the RADC cohorts, from enrollment to death, and from death to brain autopsy (Figure S3).^[1]^ We quantitatively evaluated the potential for selection bias driven by processes occurring after enrollment in the study cohorts. We also address the possibility that selection bias could have been induced by processes driving enrollment.

*Death and autopsy among those enrolled in the four RADC cohorts*. Individuals whose data were included in our study were members of any of four well-defined cohort studies: MAP, MARS, LATC and RCC. This made it possible to compare the characteristics of the 909 deceased and autopsied participants in our overall analytic sample (“Underwent autopsy” in Figure S3) with the characteristics of the larger pool of 1,276 participants who had died as of 2019 (“Died” in Figure S3), and with the characteristics of all 3,454 participants of MAP, MARS, LATC and CC (“Enrolled in cohort” in Figure S3). We first compared the demographic characteristics of these groups. On average, those in the analytic sample of autopsied participants were not markedly different in terms of age, sex, education and SES from the larger pool of participants who had died (Table S1). By contrast, Black participants comprised a smaller fraction of the autopsied group. Compared with the enrolled cohort members, those in the analytic sample were, on average, older at enrollment (mean, 82.7 versus 77.9 years; Table S1), less likely to be female (69.5% versus 75.3%), or less likely to be Black (6.5% versus 33.5%).

We then compared the three groups’ antemortem air pollution exposure, cognitive performance, and dementia status. These comparisons are tied to the principle that potential for selection bias arises when selection depends on exposure and correlates of the outcome under study, a form of collider bias.^1,2^ (Here, we used cognitive performance and dementia status as surrogates for neuropathologic load.) Mean antemortem exposures to PM_2.5_, NO_2_, and O_3_ were similar in the autopsied group and the pool of those who died (Table S1). The prevalence of dementia as of death (38.6% vs 34.3%) was slightly higher and mean global cognitive performance score was slightly lower among the former group. We also compared cognitive performance and dementia status in the autopsied sample and in all participants in the four cohorts (using their last visit). Not surprisingly, dementia was more prevalent and global cognitive scores were discernibly lower, on average, in the autopsied sample than in the cohort enrollees. These comparisons of exposure and outcome risks are unadjusted, however, and could be subject to confounding. For example, the association of dementia status with dying could be due to age.

*Air pollution exposure and neuropathologic risk in relation to selection following cohort enrollment.* Using a causal framework, we further evaluated air pollution exposure and neuropathologic risk as determinants of differential selection from cohort enrollment to autopsy. (Again, we continued to use dementia status and cognitive performance as surrogates of dementia neuropathologic risk.) The major steps in this approach were to: (1) enumerate the study population of cohort enrollees from which the analytical sample was drawn; (2) estimate the independent associations of air pollution exposure and neuropathologic risk with selection, adjusted for plausible sources of confounding; and (3) estimate the joint association of air pollution exposure and neuropathologic risk with selection, adjusted for plausible sources of confounding. The joint association was of particular interest, because simulation studies have shown that the magnitude of selection bias is much larger when exposure and outcome risk jointly determine selection (i.e., super-additively or supra-additively on the selection modeling scale) than when they determine selection independently.^3,4^

This evaluation focused only on MAP participants, because the small number of autopsies contributed by MARS (N=32), RCC (N=13), and LATC (N=1) decedents to the analytic sample posed a high risk of positivity violations in constructing the selection models. Moreover, MAP decedents comprised more than 95% of the total autopsies available for this study. For each MAP enrollee, we computed their exposure to air pollution over the past five years at baseline and at each subsequent observation cycle. Based on their enrollment date and date of death (if they had died), 2079 MAP participants—which we will call the study population—could have been in the analyses of nationally modeled PM_2.5_ had they died between 2005 and 2019 and undergone autopsy. Of this study population, 950 died between 2005 and 2019, and 791 underwent autopsy (Figure S4). Analogously for NO_2_, 2175 participants could have been in the analyses had they died between 1997 and 2019 and undergone autopsy. Of this NO_2_ study population, 1046 died, and 861 underwent autopsy (Figure S5). Note: autopsy sample sizes for this selection bias analysis differ slightly from those in our primary analysis of air pollution in relation to dementia-related neuropathology. This is because air pollution averages were anchored to visit dates in these selection bias analyses, which included both those who died and those who had not. In contrast, in the analyses of air pollution and neuropathology, the five-year average pollutant exposures were anchored to date of death.

We probed each pollutant-specific study population for three associations with the composite outcome of death and autopsy (“death+autopsy”)—specifically, the independent association of air pollution exposure, the independent association of dementia-related neuropathology risk (using a composite measure of cognitive performance as a proxy), and the joint association of air pollution and dementia-related neuropathology risk with death+autopsy.^2,3^ Notably, this composite outcome mostly indicates death, because nearly all MAP participants who died underwent autopsy.

We estimated these associations using pooled logistic regression models, with one observation per observation cycle per person through death or the end of follow-up. Exposure to air pollution over the past five years was update at each cycle, as describe; most recent cognitive score was similarly updated, as well. To inform the covariate sets, we hewed to a causally informed framework, rather than a prediction framework. Thus, we fit separate models for each of the three types of association, adjusting for potential sources of confounding of each. All analyses of were adjusted for calendar year of enrollment, education (years), sex and age at visit. Analyses of joint associations additionally included terms for both air pollution and cognitive performance and the interaction between the two. We fitted separate models for each pollutant. In secondary analyses, we used air pollution exposure and cognitive performance at baseline only, following the approach used in ACT autopsy studies.^5,6,7^

Cognitive performance was strongly and inversely associated with death and autopsy, an expected result given the established association of cognitive functioning with mortality.^8-13^ For example, in the PM_2.5_ selection sample, the odds ratio (OR) of death+autopsy per SD higher global cognitive score *at most recent visit* was 0.50 (95% CI: 0.46, 0.54; Table S2). In secondary analyses of cognitive performance at baseline, 1 SD higher *baseline* global cognitive score corresponded to a death+autopsy OR of 0.84 (95% CI: 0.69, 1.03; Table S2).

By contrast, although higher exposure to air pollution is associated with mortality in the general population,^9-11^ there were no such associations in this study population with death+autopsy. For example, most recent 5-year average PM_2.5_ was inversely associated with death+autopsy [OR per µg/m^3^, 0.91 (95% CI: 0.86, 0.96)], and there was little association of most recent 5-year average NO_2_ with death+autopsy [OR per ppb, 0.97 (95% CI 0.92, 1.02) (Table S2)]. Moreover, there was little indication of any joint association involving both air pollution exposure and cognitive performance (Table S2).

This evaluation generated little evidence to suggest that our estimated associations of air pollution with dementia-related pathology were strongly influenced by selective attrition *after enrollment*, of a type that followed a classic collider bias pattern (also known as “Type 1” selection bias). It remains possible, however, that our estimates were biased by a different type of selection pattern whereby selection depended on one or more modifiers of the exposure-neuropathology effect (also called “Type 2” selection bias).^1,14,15^

*Cohort enrollment among the target population*. Even if it were true that differential selection following enrollment was minimal, other sources of selection bias could have been present. Air pollutant concentrations in the study area often exceeded levels at which associations with mortality are typically observed.^16,17^ The lack of an adverse association in the study population between air pollution exposure and mortality suggests that MAP participants, on the whole, may have been unusually robust to the adverse health effects of exposure compared with the broader population of older adults in the United States. Thus, it is possible that the enrollment process resulted in a study sample in which the effect of air pollution exposure on dementia-related pathology differed from that in this target population of older adults.^1^ Quantifying “pre-enrollment selection bias,” unfortunately, is far more challenging than “post-enrollment selection bias,” because few of the required data are available. Moreover, as newly described by Morenz (2023),^18^ estimating effects on autopsy-based outcomes not only involves the challenge of differential death, but also differential timing of death. If neuropathologic load tends to increase with time, and if exposure promotes neuropathology but also hastens death, the comparisons of neuropathology on autopsy could underestimate the effect of exposure.

**Figure S4**. Flowchart of MAP participants included in analyses of five-year average PM_2.5_ and neuropathology originating from the study population all MAP participants who could have been included had they died between 2005 and 2019 and undergone autopsy.

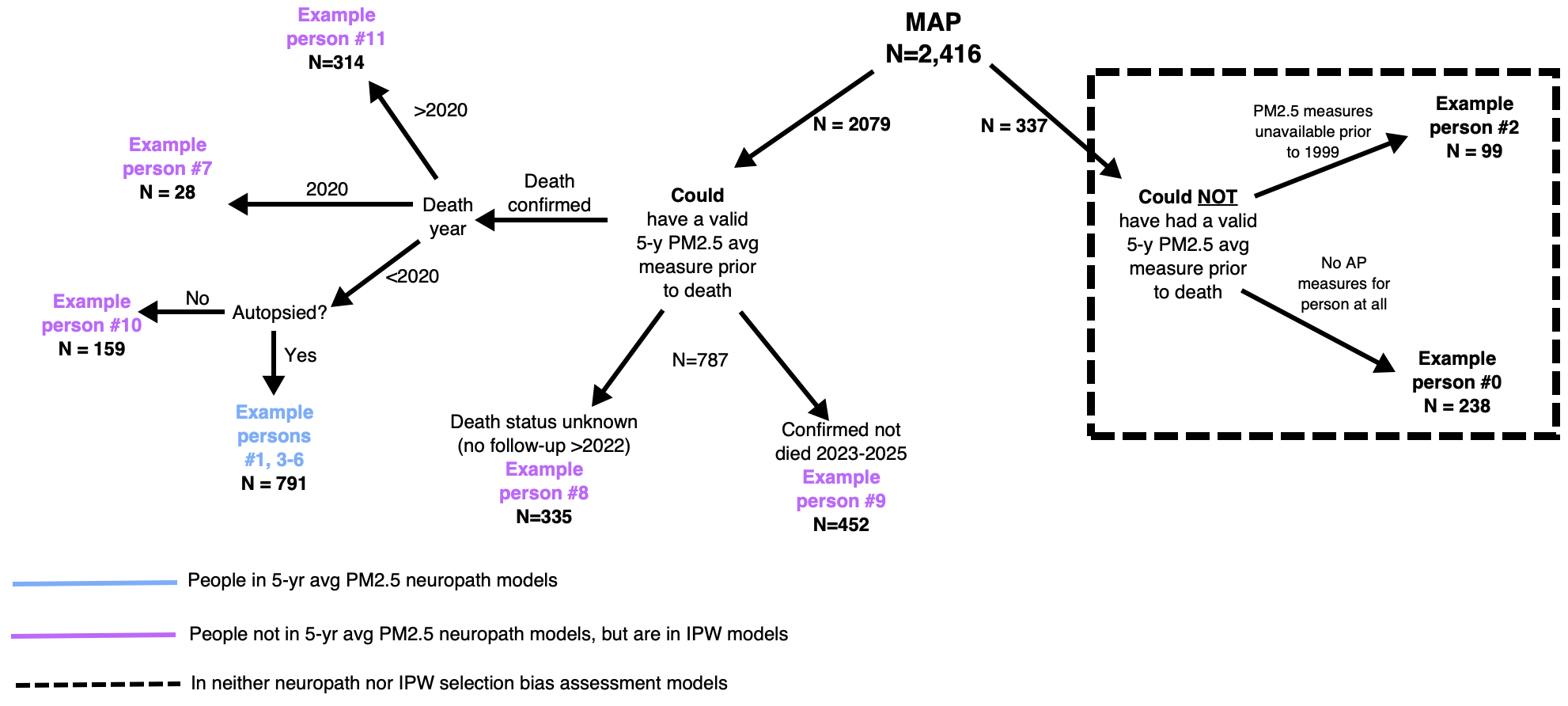

**Figure S5**. Flowchart of MAP participants included in analyses of five-year average NO_2_ and neuropathology originating from the study population all MAP participants who could have been included had they died between 1997 and 2019 and undergone autopsy.

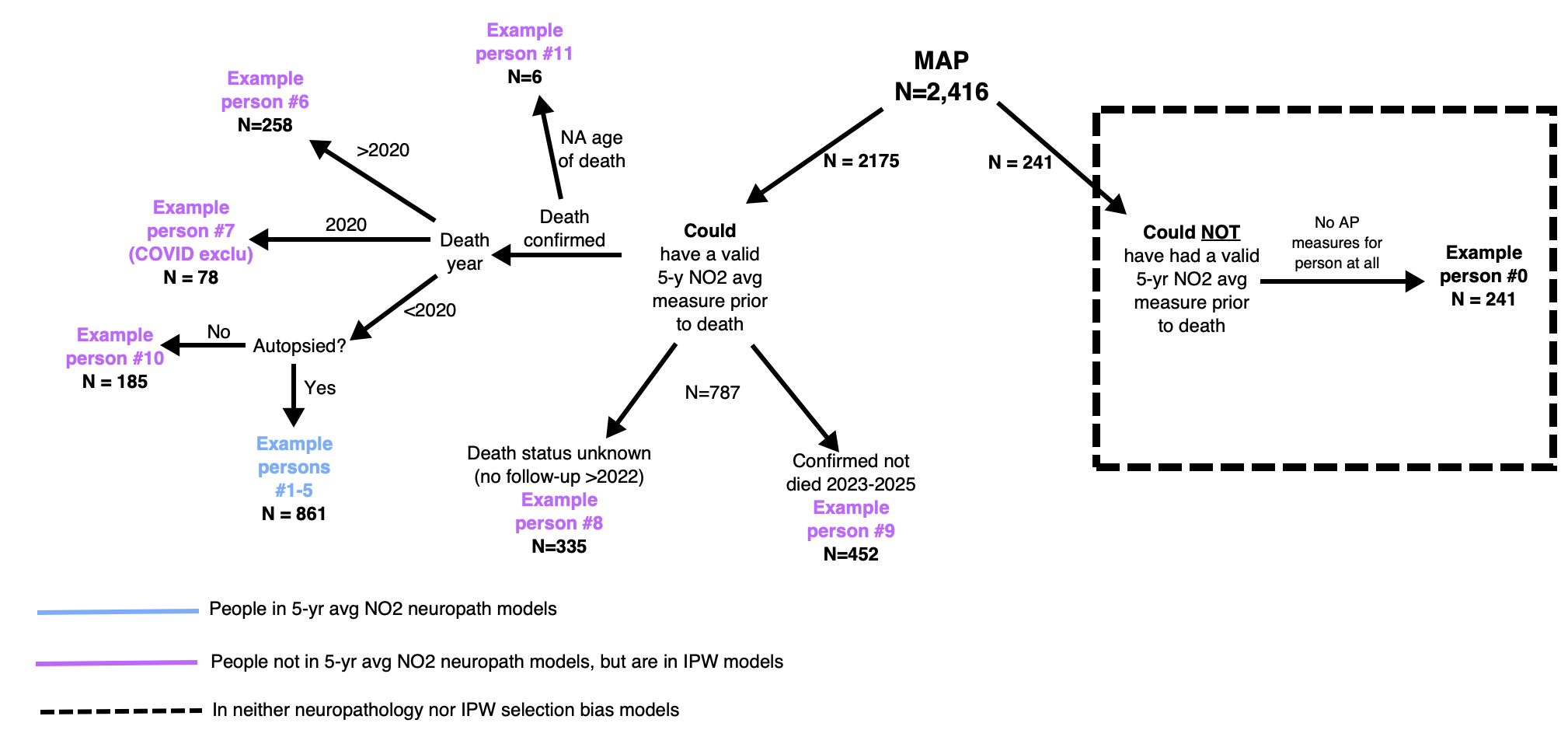

Table S1. Characteristics of combined Rush MAP, MARS, RCC, and LATC cohorts, deceased cohort and autopsy cohort.

|  | **MAP, MARS, LC & CC cohorts combined** | | |
| --- | --- | --- | --- |
|  | Total^a^ | Died^b^ | Autopsy |
|  | N=3,454 | N=1,276 | N=909 |
|  | Mean (SD)/N (%) | Mean (SD)/N (%) | Mean (SD)/N (%) |
| **Ages/Years** |  |  |  |
| Age at Enrollment (Years) (mean SD) | 77.4 (7.9) | 81.6 (6.9) | 82.7 (6.3) |
| Birth Year (mean SD) | 1932 (10.8) | 1923 (8.4) | 1922 (7.6) |
| Age at Death (Years) (mean SD)^c^ | 88.8 (7.4) | 88.5 (7.3) | 90.0 (6.6) |
| <85 years | 453 (25.3) | 336 (26.3) | 167 (18.4) |
| 85 - 89 years | 436 (24.3) | 313 (24.5) | 225 (24.8) |
| 90 - 94 years | 503 (28.1) | 373 (29.2) | 299 (32.9) |
| 95+ years | 401 (22.4) | 254 (19.9) | 218 (24.0) |
| Year of Death (mean SD)^c^ | 2015 (6.4) | 2012 (5.0) | 2012 (5.0) |
| **Demographics** |  |  |  |
| Sex |  |  |  |
| Female | 2601 (75.3) | 882 (69.1) | 632 (69.5) |
| Male | 853 (24.7) | 394 (30.9) | 277 (30.5) |
| Race/ethnicity |  |  |  |
| Black | 1157 (33.5) | 259 (20.3) | 59 (6.5) |
| Other | 190 (5.5) | 11 (0.9) | 3 (0.3) |
| White | 2107 (61.0) | 1006 (78.8) | 847 (93.2) |
| Spanish/Hispanic/Latino origin |  |  |  |
| Hispanic | 359 (10.4) | 30 (2.4) | 17 (1.9) |
| Non-Hispanic | 3095 (89.6) | 1246 (97.6) | 892 (98.1) |
| Missing | 1 (0.0) | - | - |
| Education |  |  |  |
| Less than High School | 352 (10.2) | 106 (8.3) | 55 (6.1) |
| High School | 712 (20.6) | 312 (24.5) | 226 (24.9) |
| More than High School | 2390 (69.2) | 858 (67.2) | 628 (69.1) |
| Early-Life SES |  |  |  |
| Lowest Quartile | 778 (22.5) | 299 (23.4) | 220 (24.2) |
| Quartile 2 | 778 (22.5) | 345 (27.0) | 220 (24.2) |
| Quartile 3 | 777 (22.5) | 296 (23.2) | 220 (24.2) |
| Highest Quartile | 777 (22.5) | 265 (20.8) | 219 (24.1) |
| Missing | 334 (10.0) | 71 (5.6) | 30 (3.3) |
| Income at Age 40 |  |  |  |
| <$30,000 | 1448 (41.9) | 623 (48.8) | 438 (48.2) |
| $30,000 - $49,999 | 960 (27.8) | 286 (22.4) | 195 (21.5) |
| $50,000 - $74,999 | 421 (12.2) | 87 (6.8) | 56 (6.2) |
| $75,000+ | 223 (6.5) | 41 (3.2) | 25 (2.8) |
| Missing | 402 (11.6) | 239 (18.7) | 195 (21.5) |
| Baseline Income |  |  |  |
| <$30,000 | 1307 (37.8) | 477 (37.4) | 296 (32.6) |
| $30,000 - $49,999 | 815 (23.6) | 300 (23.5) | 221 (24.3) |
| $50,000 - $74,999 | 504 (14.6) | 157 (12.3) | 119 (13.1) |
| $75,000+ | 545 (15.8) | 144 (11.3) | 112 (12.3) |
| Missing | 283 (8.2) | 198 (15.5) | 161 (17.7) |
| Smoking status |  |  |  |
| Never | 1969 (57.0) | 707 (55.4) | 537 (59.1) |
| Former | 1312 (38.0) | 499 (39.1) | 340 (37.4) |
| Current | 153 (4.4) | 59 (4.6) | 25 (2.8) |
| Missing | 20 (0.6) | 11 (0.9) | 7 (0.8) |
| Apolipoprotein E genetics |  |  |  |
| ApoE4 (at least 1) | 783 (22.7) | 308 (24.1) | 215 (23.7) |
| No ApoE4 alleles | 2033 (58.9) | 917 (71.9) | 694 (73.7) |
| Missing | 638 (18.5) | 51 (4.0) | 24 (2.6) |
| **Cognitive status at last visit** |  |  |  |
| Dementia diagnosis - N (%) |  |  |  |
| Yes | 797 (23.1) | 438 (34.3) | 351 (38.6) |
| No | 2348 (68.0) | 803 (62.9) | 533 (58.6) |
| Missing | 309 (8.9) | 35 (2.7) | 25 (2.8) |
| Cognition - Mean (SD) |  |  |  |
| Global cognitive z-score | -0.49 (0.96) | -0.77 (1.01) | -0.81 (1.05) |
| Missing (N) | (377) | (147) | (117) |
| **5-year air pollution concentration Mean (SD)** |  |  |  |
| National PM_2.5_ ug/m^3^ | * | 10.27 (1.6) | 10.1 (1.6) |
| Missing pollution measure (N) | * | (98) | (72) |
| National NO_2_ ppb | * | 12.39 (4.5) | 11.4 (4.1) |
| Missing pollution measure (N) | * | (2) | - |
| National 'O_3_ ppb | * | 22.6 (2.3) | 23.0 (2.1) |
| Missing pollution measure (N) | * | (2) | - |

^a^Rush MAP, MARS, LATC, and RCC participants with 5-year prior average NO2/O3 air pollution measures at baseline

^b^Subset of participants with air pollution measures at baseline who died after enrollment and prior to 2020

^c^1661 participants still alive at time of analysis

*Pollutant measures prior to death available only for sample who have died

Table S2. Adjusted associations of cognitive score and pollutant exposure with death followed by autopsy, among MAP participants.

|  |  | **PM2.5 sample (N=2079)** | | |  | | **NO2 sample (N=2175)** | | |
| --- | --- | --- | --- | --- | --- | --- | --- | --- | --- |
|  |  | **Odds ratio** | **(95% confidence interval)** | |  | **Odds ratio** | | **(95% confidence interval)** | |
| *MAIN EFFECTS ESTIMATION* | |  |  |  |  |  | |  |  |
|  | *Time-updated* |  |  |  |  |  | |  |  |
|  | Most recent cognitive score, per SD unit* | 0.50 | (0.46, 0.54) | |  | 0.51 | | (0.47, 0.54) | |
|  | *Baseline* |  |  |  |  |  | |  |  |
|  | Baseline cognitive score, per SD unit** | 0.84 | (0.69, 1.03) | |  | 0.98 | | (0.84, 1.16) | |
|  | *Time-updated* |  |  |  |  |  | |  |  |
|  | PM2.5 exposure in the past 5 years, per ug/m3* | 0.91 | (0.86, 0.96) | |  | - | | - | - |
|  | NO2 exposure in the past 5 years, per ppb* | - | - | - |  | 0.97 | | (0.92, 1.02) | |
|  | *Baseline* |  |  |  |  |  | |  |  |
|  | PM2.5 exposure in the 5 years before baseline, per ug/m3* | 0.78 | (0.66, 0.92) | |  | - | | - | - |
|  | NO2 exposure in the 5 years before baseline, per ppb* | - | - | - |  | 0.96 | | (0.94, 0.98) | |
| *JOINT EFFECTS ESTIMATION* | |  |  |  |  |  | |  |  |
|  | *Time-updated**** |  |  |  |  |  | |  |  |
|  | Most recent cognitive score, per SD unit | 0.50 | (0.46, 0.54) | |  | - | | - | - |
|  | PM2.5 exposure in the past 5 years, per ug/m3 | 0.90 | (0.84, 0.97) | |  | - | | - | - |
|  | Interaction | 1.04 | (0.99, 1.09) | |  | - | | - | - |
|  | Most recent cognitive score, per SD unit | - | - | - |  | 0.51 | | (0.47, 0.56) | |
|  | NO2 exposure in the past 5 years, per ppb | - | - | - |  | 0.97 | | (0.95, 1.00) | |
|  | Interaction | - | - | - |  | 1.01 | | (0.99, 1.03) | |
|  | *Baseline***** |  |  |  |  |  | |  |  |
|  | Baseline cognitive score, per SD unit | 0.78 | (0.63, 0.95) | |  | - | | - | - |
|  | PM2.5 exposure in the 5 years before baseline, per ug/m3* | 0.80 | (0.68, 0.95) | |  | - | | - | - |
|  | Interaction | 1.30 | (1.17, 1.44) | |  | - | | - | - |
|  | Baseline cognitive score, per SD unit | - | - | - |  | 0.91 | | (0.77, 1.08) | |
|  | NO2 exposure in the 5 years before baseline, per ppb* | - | - | - |  | 0.97 | | (0.95, 0.99) | |
|  | Interaction | - | - | - |  | 1.07 | | (1.04, 1.10) | |

* Based on pooled logistic regression models and adjusted for current age, current calendar year, sex, and education. Note: this odds ratio can be interpreted as a hazard ratio.

** Based on logistic regression models and adjusted for baseline age, baseline calendar year, sex, and education.

*** Based on pollutant-specific pooled regression models that included cognitive score, the pollutant, and the interaction between the two. Analyses were adjusted for current age, current calendar year, sex, and education.

**** Based on pollutant-specific logistic regression models that included cognitive score, the pollutant, and the interaction between the two. Analyses were adjusted for baseline age, baseline calendar year, sex, and education.

**Figure S6**. Construction of Two Year-Lag Average Pollutant Exposures Prior to Death for Sensitivity Models. RADC Cohorts: MAP, MARS, LATC, RCC (predicted by Chicago-specific model, N=775; predicted by national models, N=837)

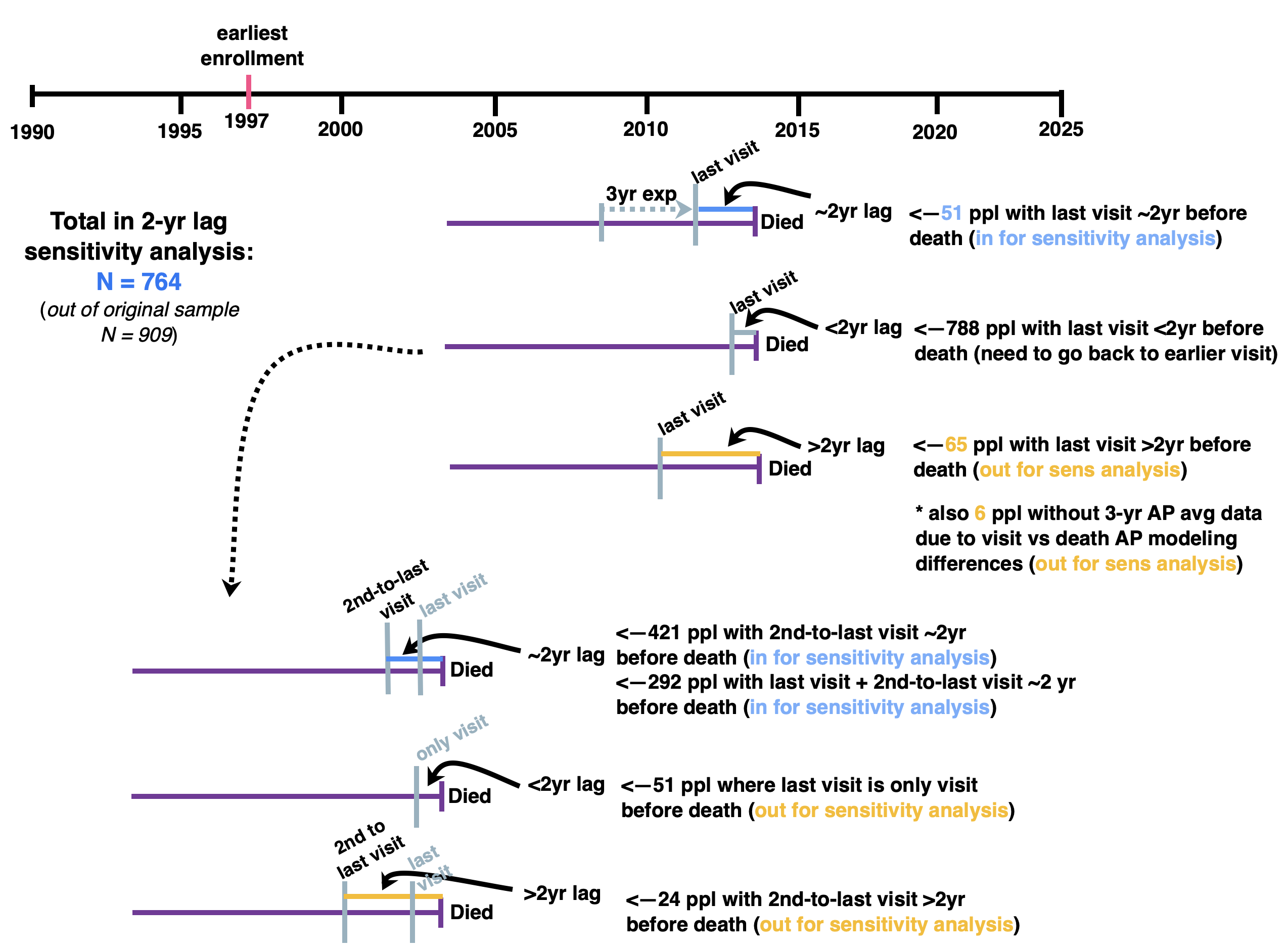

**Figure S7**. Five-Year Average PM_2.5_ Prior to Death, by Year of Death. RADC Cohorts: MAP, MARS, LATC, RCC (predicted by Chicago-specific model, N=775; predicted by national models, N=837)

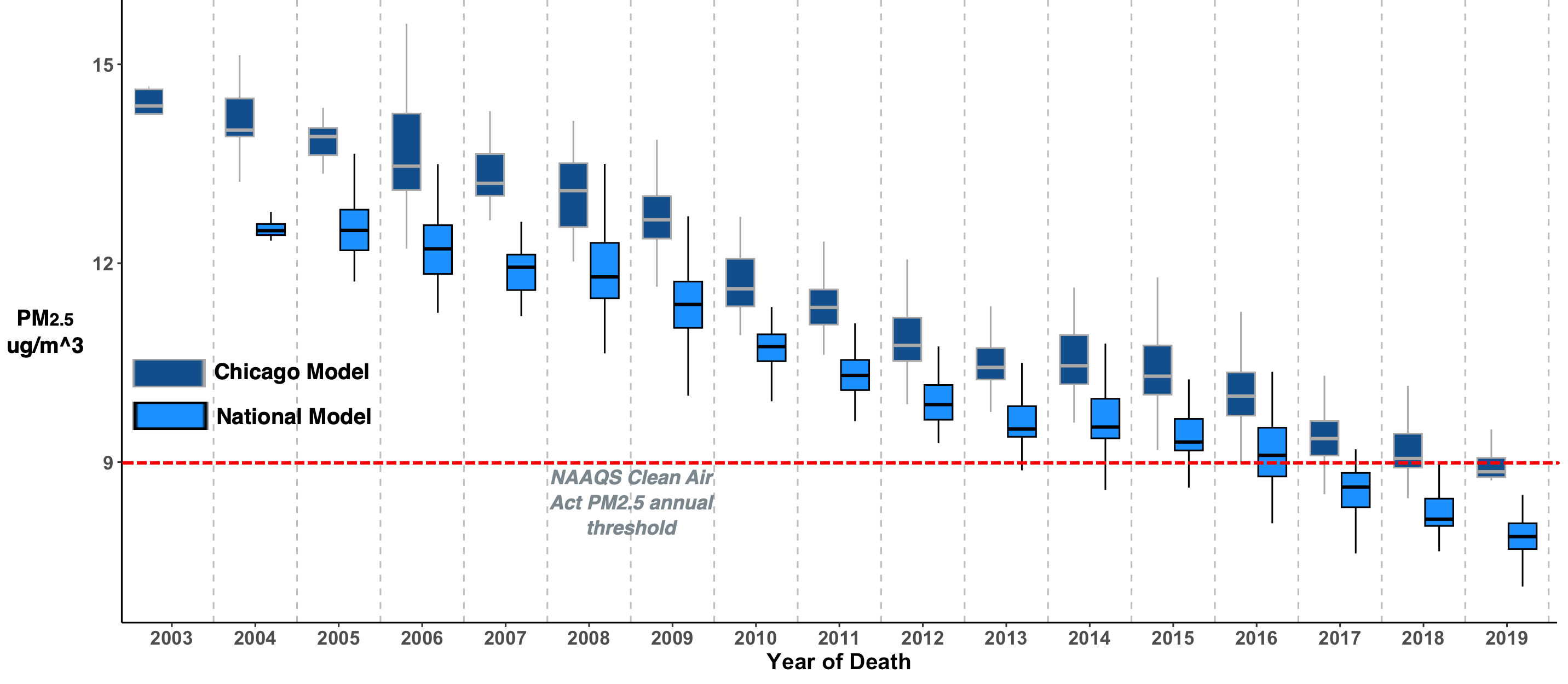

**Figure S8.** Five-Year Average NO_2_ and NO_x_ (Chicago and National Models) Prior to Death, by Year of Death. RADC Cohorts: MAP, MARS, LATC, RCC (N=909 National Models, N=775 Chicago Models )

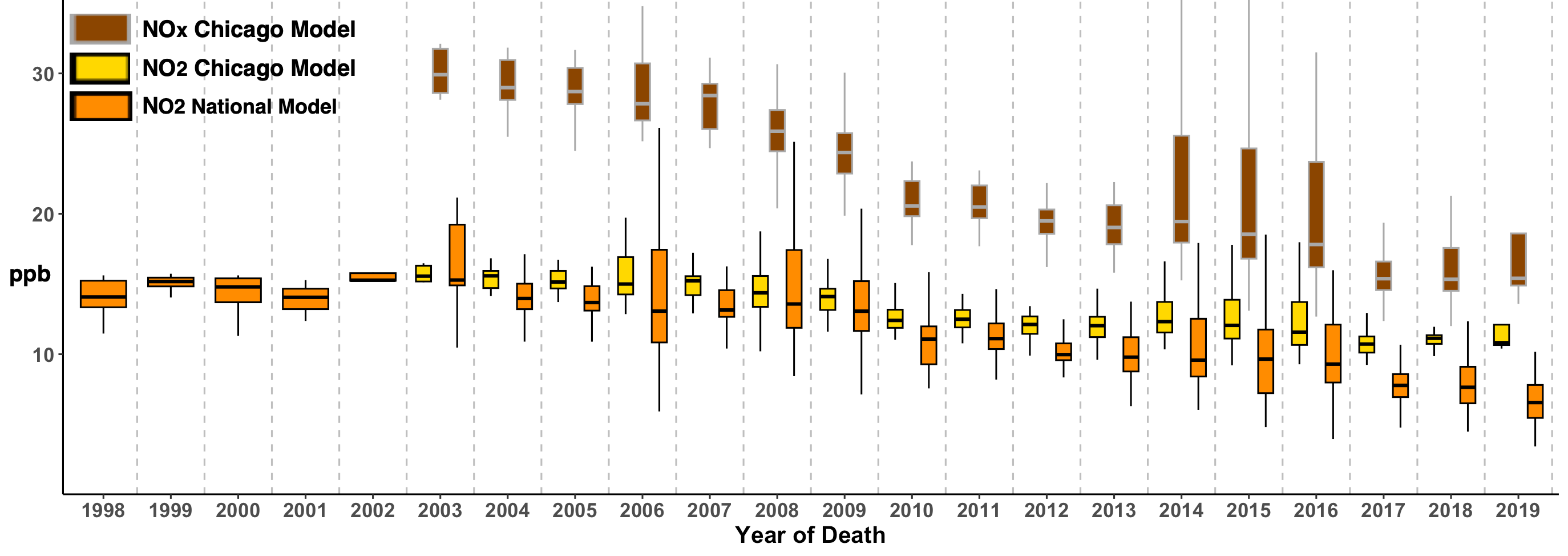

**Figure S9**. Five-Year Average O_3_ in National and Chicago Models Prior to Death, by Year of Death. RADC Cohorts: MAP, MARS, LATC, RCC (National models: N=909, Chicago models N=749)

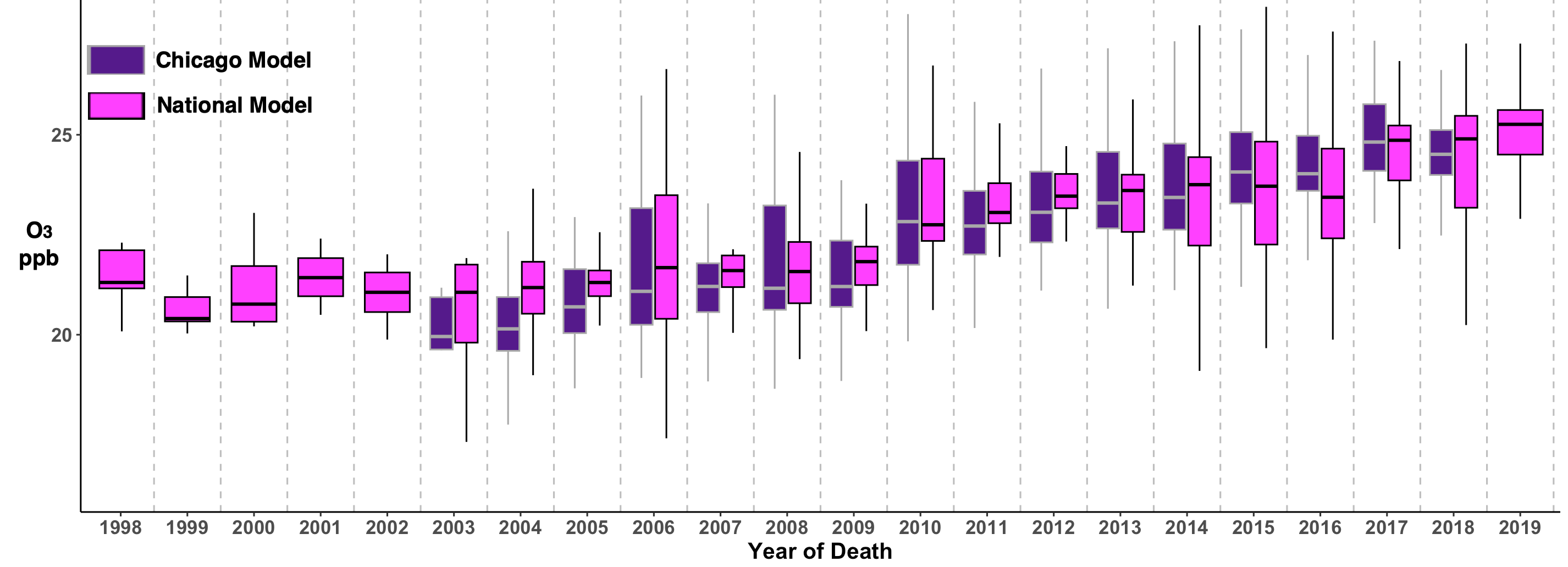

**Table S3**. Estimated mean difference and 95% confidence intervals (CI) in beta-amyloid density and tau tangle density at autopsy per increment in 5-year average exposure to national- and Chicago-modeled air pollutants (see Figure 2). RADC Cohorts: MAP, MARS, LATC, RCC.

|  | Mean difference (95% CI) | |
| --- | --- | --- |
|  | Beta-amyloid density | Tau tangle  density |
| Nat'l PM2.5 Five-Year Average  Exposure - per 2.5 ug/m3 increase* | -0.25 (-0.47, -0.02) | 0.09 (-0.20, 0.39) |
| Chicago PM2.5 Five-Year Average Exposure - per 2.5 ug/m3 increase** | -0.39 (-0.63, -0.15) | 0.25 (-0.05, 0.56) |
| Nat'l NO2 Five-Year Average**  Exposure - per 5 ppb increase | -0.04 (-0.12, 0.05) | 0.02 (-0.09, 0.12) |
| Chicago NO2 Five-Year Average** Exposure - per 5 ppb increase | -0.16 (-0.30, -0.03) | 0.10 (-0.07, 0.28) |
| Chicago NOx Five-Year Average** Exposure - per 5 ppb increase | -0.11 (-0.18, -0.04) | 0.04 (-0.05, 0.12) |
| Nat'l O3 Five-Year Average***  Exposure - per 2.5 ppb increase | 0.01 (-0.06, 0.09) | -0.03 (-0.13, 0.07) |
| Chicago O3 Five-Year Average**** Exposure - per 2.5 ppb increase | 0.07 (-0.02, 0.15) | -0.04 (-0.15, 0.06) |
| *Total N = 837 (Nat'l PM2.5) | N=836 | N=834 |
| **Total N = 775 (Chi PM2.5/NO2/NOx) | N=774 | N=772 |
| *** N = 909 (Nat'l NO2/O3) | N=908 | N=906 |
| ****N = 749 Chi O3 | N=748 | N=746 |

Exposures computed over the 5 years prior to death, using national and Chicago prediction models. N = sample size for continuous outcomes. Beta-amyloid (Aβ) density was measured as the square root-transformed mean percent of Aβ positivity in the cortex. Tau tangle density was measured as square root-transformed mean of tau-tangles per square mm. We used linear regression models to estimate the mean difference per exposure increment in β-amyloid and tau tangle densities. Models were adjusted for time to autopsy, birth year, death year, sex, race/ethnicity, years of education, baseline smoking status, baseline income, income at age 40, early life socioeconomic status, ≥ 1 ApoE4 allele. AD: Alzheimer’s disease. RADC: Rush Alzheimer’s Disease Center. MAP: Memory and Aging Project. MARS: Minority Aging Research Study. LATC: Latino Core, RCC: Rush Clinical Core.

**Table S4.** Risk ratios (RR) and 95% confidence intervals (CI) for association of 5-year average exposure to national- and Chicago-modeled air pollutants with ADNC, cerebral arteriolosclerosis, cerebral atherosclerosis, and cerebral amyloid angiography at autopsy (see Figures 2 and 3). RADC Cohorts: MAP, MARS, LATC, RCC.

|  | ADNC (AD present/not) | Cerebral arteriolosclerosis  (none/mild vs moderate/severe) | Cerebral  atherosclerosis (none/mild vs moderate/severe) | Cerebral amyloid  angiopathy (none/mild vs moderate/severe) |
| --- | --- | --- | --- | --- |
|  | RR (95% CI) | RR (95% CI) | RR (95% CI) | RR (95% CI) |
| Nat'l PM2.5 Five-Year Average Exposure - per 2.5 ug/m3 increase* | 0.94 (0.74, 1.17) | 1.41 (0.95, 2.09) | 1.21 (0.80, 1.81) | 1.04 (0.68, 1.58) |
| Chicago PM2.5 Five-Year Average Exposure - per 2.5 ug/m3 increase** | 0.96 (0.76, 1.20) | 1.51 (1.02, 2.24) | 1.41 (0.93, 2.13) | 1.00 (0.64, 1.55) |
| Nat'l NO2 Five-Year Average Exposure - per 5 ppb increase*** | 0.99 (0.92, 1.08) | 1.07 (0.95, 1.21) | 0.98 (0.84, 1.14) | 1.02 (0.88, 1.19) |
| Chicago NO2 Five-Year Average Exposure - per 5 ppb increase** | 0.98 (0.86, 1.13) | 1.07 (0.85, 1.34) | 1.06 (0.84, 1.35) | 1.05 (0.82, 1.34) |
| Chicago NOx Five-Year Average Exposure - per 5 ppb increase** | 1.00 (0.93, 1.07) | 1.09 (0.98, 1.22) | 1.10 (0.98, 1.23) | 0.97 (0.85, 1.10) |
| Nat'l O3 Five-Year Average Exposure - per 2.5 ug/m3 increase*** | 1.00 (0.93, 1.07) | 0.88 (0.76, 1.01) | 0.96 (0.82, 1.12) | 1.02 (0.89, 1.17) |
| Chicago O3 Five-Year Average Exposure - per 2.5 ug/m3 increase**** | 1.04 (0.96, 1.12) | 0.85 (0.73, 0.99) | 0.95 (0.82, 1.10) | 1.05 (0.91, 1.23) |
| *Total N = 837 (Nat'l PM2.5) | N=542/837 | N=266/835 | N=232/836 | N=288/835 |
| **Total N = 775 (Chi PM2.5/NO2/NOx) | N=507/775 | N=252/773 | N=221/774 | N=268/772 |
| ***Total N = 909 (Nat'l NO2/O3) | N=586/909 | N=305/907 | N=266/908 | N=315/903 |
| ****Total N = 749 (Chi O3) | N=493/749 | N=250/747 | N=217/748 | N=257/746 |

Exposures computed over the 5 years prior to death, using national and Chicago prediction models. N = events/total sample size for dichotomous outcomes. We used log binomial regression models to estimate prevalence ratios per exposure increment of ADNC, cerebral arteriolosclerosis, cerebral atherosclerosis, cerebral amyloid angiopathy, vascular chronic infarcts (gross, microscopic, and total), Lewy bodies, hippocampal sclerosis, and LATE-NC. Models were adjusted for time to autopsy, birth year, death year, sex, race/ethnicity, years of education, baseline smoking status, baseline income, income at age 40, early life socioeconomic status, ≥ 1 ApoE4 allele. ADNC: Alzheimer’s disease neuropathology. RADC: Rush Alzheimer’s Disease Center. MAP: Memory and Aging Project. MARS: Minority Aging Research Study. LATC: Latino Core, RCC: Rush Clinical Core.

**Table S5.** Risk ratios (RR) and 95% confidence intervals (CI) for association of 5-year average exposure to national- and Chicago-modeled air pollutants with chronic cerebrovascular infarctions at autopsy (See Figure 3). RADC Cohorts: MAP, MARS, LATC, RCC.

|  | Gross chronic infarcts (none vs any) | Microscopic chronic infarcts (none vs any) | Total gross or microscopic chronic infarcts (none vs any) |
| --- | --- | --- | --- |
|  | RR (95% CI) | RR (95% CI) | RR (95% CI) |
| Nat'l PM2.5 Five-Year Average Exposure - per 2.5 ug/m3 increase* | 0.70 (0.47, 1.07) | 0.65 (0.40, 1.06) | 0.89 (0.66, 1.21) |
| Chicago PM2.5 Five-Year Average Exposure - per 2.5 ug/m3 increase** | 0.68 (0.44, 1.05) | 0.91 (0.56, 1.48) | 0.90 (0.66, 1.24) |
| Nat'l NO2 Five-Year Average Exposure - per 5 ppb increase*** | 0.87 (0.75, 1.02) | 0.96 (0.80, 1.16) | 0.96 (0.85, 1.08) |
| Chicago NO2 Five-Year Average Exposure - per 5 ppb increase** | 0.81 (0.62, 1.06) | 0.97 (0.73, 1.29) | 0.96 (0.80, 1.17) |
| Chicago NOx Five-Year Average Exposure - per 5 ppb increase** | 0.93 (0.81, 1.05) | 0.97 (0.85, 1.12) | 0.99 (0.90, 1.09) |
| Nat'l O3 Five-Year Average Exposure - per 2.5 ug/m3 increase*** | 1.13 (1.00, 1.28) | 1.03 (0.88, 1.21) | 1.02 (0.93, 1.13) |
| Chicago O3 Five-Year Average Exposure - per 2.5 ug/m3 increase**** | 1.11 (0.97, 1.28) | 0.97 (0.82, 1.15) | 1.05 (0.94, 1.16) |
| *Total N = 837 (Nat'l PM2.5) | N=313/837 | N=263/837 | N=435/837 |
| **Total N = 775 (Chi PM2.5/NO2/NOx) | N=289/775 | N=237/775 | N=402/775 |
| ***Total N = 909 (Nat'l NO2/O3) | N=332/909 | N=274/909 | N=461/909 |
| ****Total N = 749 (Chi O3) | N=281/749 | N=227/749 | N=387/749 |

Exposures computed over the 5 years prior to death, using national and Chicago prediction models. N = events/total sample size for dichotomous outcomes. We used log binomial regression models to estimate prevalence ratios per exposure increment of chronic infarcts (gross, microscopic, and total). Models were adjusted for time to autopsy, birth year, death year, sex, race/ethnicity, years of education, baseline smoking status, baseline income, income at age 40, early life socioeconomic status, ≥ 1 ApoE4 allele. AD: Alzheimer’s disease. RADC: Rush Alzheimer’s Disease Center. MAP: Memory and Aging Project. MARS: Minority Aging Research Study. LATC: Latino Core, RCC: Rush Clinical Core.

**Table S6.** Risk ratios (RR) and 95% confidence intervals for association of 5-year average exposure to national- and Chicago-modeled air pollutants with Lewy bodies, hippocampal sclerosis, and LATE-NC at autopsy (see Figure 4). RADC Cohorts: MAP, MARS, LATC, RCC.

|  | Lewy bodies  (absent vs present) | Hippocampal sclerosis  (absent vs present) | LATE-NC (stage 0/1 vs stage 2/3) |
| --- | --- | --- | --- |
|  | RR (95% CI) | RR (95% CI) | RR (95% CI) |
| Nat'l PM2.5 Five-Year Average Exposure - per 2.5 ug/m3 increase* | 0.93 (0.54, 1.59) | 0.52 (0.20, 1.38) | 0.95 (0.62, 1.45) |
| Chicago PM2.5 Five-Year Average Exposure - per 2.5 ug/m3 increase** | 0.69 (0.40, 1.20) | 0.34 (0.12, 0.95) | 1.22 (0.78, 1.90) |
| Nat'l NO2 Five-Year Average Exposure - per 5 ppb increase*** | 0.96 (0.79, 1.16) | 0.89 (0.61, 1.29) | 1.02 (0.86, 1.20) |
| Chicago NO2 Five-Year Average Exposure - per 5 ppb increase** | 0.79 (0.57, 1.11) | 0.70 (0.35, 1.37) | 1.05 (0.81, 1.37) |
| Chicago NOx Five-Year Average Exposure - per 5 ppb increase** | 0.90 (0.77, 1.06) | 0.75 (0.51, 1.09) | 1.01 (0.89, 1.15) |
| Nat'l O3 Five-Year Average Exposure - per 2.5 ug/m3 increase*** | 1.08 (0.90, 1.28) | 1.13 (0.85, 1.50) | 0.99 (0.86, 1.14) |
| Chicago O3 Five-Year Average Exposure - per 2.5 ug/m3 increase**** | 1.09 (0.90, 1.32) | 1.13 (0.83, 1.53) | 0.95 (0.81, 1.11) |
| *Total N = 837 (Nat'l PM2.5) | N=226/837 | N=89/837 | N=301/829 |
| **Total N = 775 (Chi PM2.5/NO2/NOx) | N=203/775 | N=79/775 | N=267/767 |
| ***Total N = 909 (Nat'l NO2/O3) | N=235/909 | N=90/908 | N=308/885 |
| ****Total N = 749 (Chi O3) | N=196/749 | N=77/749 | N=257/741 |

Exposures computed over the 5 years prior to death, using national and Chicago prediction models. N = events/total sample size for dichotomous outcomes. We used log binomial regression models to estimate prevalence ratios per exposure increment of Lewy bodies, hippocampal sclerosis, and LATE-NC. Models were adjusted for time to autopsy, birth year, death year, sex, race/ethnicity, years of education, baseline smoking status, baseline income, income at age 40, early life socioeconomic status, ≥ 1 ApoE4 allele. RADC: Rush Alzheimer’s Disease Center. MAP: Memory and Aging Project. MARS: Minority Aging Research Study. LATC: Latino Core, RCC: Rush Clinical Core. LATE-NC: limbic predominant age-related TDP-43 encephalopathy - neuropathologic change.

**Table S7**. Results from sensitivity analysis (a). Estimated mean difference and 95% confidence intervals (CI) in beta-amyloid density and tau tangle density at autopsy per increment in 5-year average exposure to national- and Chicago-modeled air pollutants, restricted to the subset of 730 participants for whom we were able to estimate all seven national-modeled and Chicago-modeled five-year average exposures. RADC Cohorts: MAP, MARS, LATC, RCC.

|  | Mean difference (95% CI) | | |
| --- | --- | --- | --- |
|  | Beta-amyloid density | Tau tangle  density | |
| Nat'l PM2.5 Five-Year Average  Exposure - per 2.5 ug/m3 increase | -0.24 (-0.49, 0.003) | | 0.15 (-0.16, 0.46) |
| Chicago PM2.5 Five-Year Average Exposure - per 2.5 ug/m3 increase | -0.41 (-0.65, -0.17) | | 0.20 (-0.10, 0.50) |
| Nat'l NO2 Five-Year Average Exposure - per 5 ppb increase | -0.05 (-0.15, 0.04) | | 0.05 (-0.07, 0.17) |
| Chicago NO2 Five-Year Average Exposure - per 5 ppb increase | -0.18 (-0.32, -0.03) | | 0.11 (-0.08, 0.29) |
| Chicago NOx Five-Year Average Exposure - per 5 ppb increase | -0.12 (-0.19, -0.05) | | 0.03 (-0.05, 0.12) |
| Nat'l O3 Five-Year Average Exposure - per 2.5 ppb increase | 0.06 (-0.03, 0.16) | | -0.06 (-0.18, 0.06) |
| Chicago O3 Five-Year Average Exposure - per 2.5 ppb increase | 0.07 (-0.01, 0.16) | | -0.03 (-0.14, 0.08) |
| N = 730 | N=729 | | N=727 |

Exposures computed over the 5 years prior to death, using national and Chicago prediction models. N = sample size for continuous outcomes. Beta-amyloid (Aβ) density was measured as the square root-transformed mean percent of Aβ positivity in the cortex. Tau tangle density was measured as square root-transformed mean of tau-tangles per square mm. We used linear regression models to estimate the mean difference per exposure increment in β-amyloid and tau tangle densities. Models were adjusted for time to autopsy, birth year, death year, sex, race/ethnicity, years of education, baseline smoking status, baseline income, income at age 40, early life socioeconomic status, ≥ 1 ApoE4 allele. AD: Alzheimer’s disease. RADC: Rush Alzheimer’s Disease Center. MAP: Memory and Aging Project. MARS: Minority Aging Research Study. LATC: Latino Core, RCC: Rush Clinical Core.

**Table S8**. Results from sensitivity analysis (a). Risk ratios (RR) and 95% confidence intervals (CI) for association of 5-year average exposure to national- and Chicago-modeled air pollutants with ADNC, cerebral arteriolosclerosis, cerebral atherosclerosis, cerebral amyloid angiography, chronic cerebrovascular infarctions, Lewy bodies, hippocampal sclerosis, and LATE-NC at autopsy, restricted to the subset of 730 participants for whom we were able to estimate all seven national-modeled and Chicago-modeled five-year average exposures. RADC Cohorts: MAP, MARS, LATC, RCC.

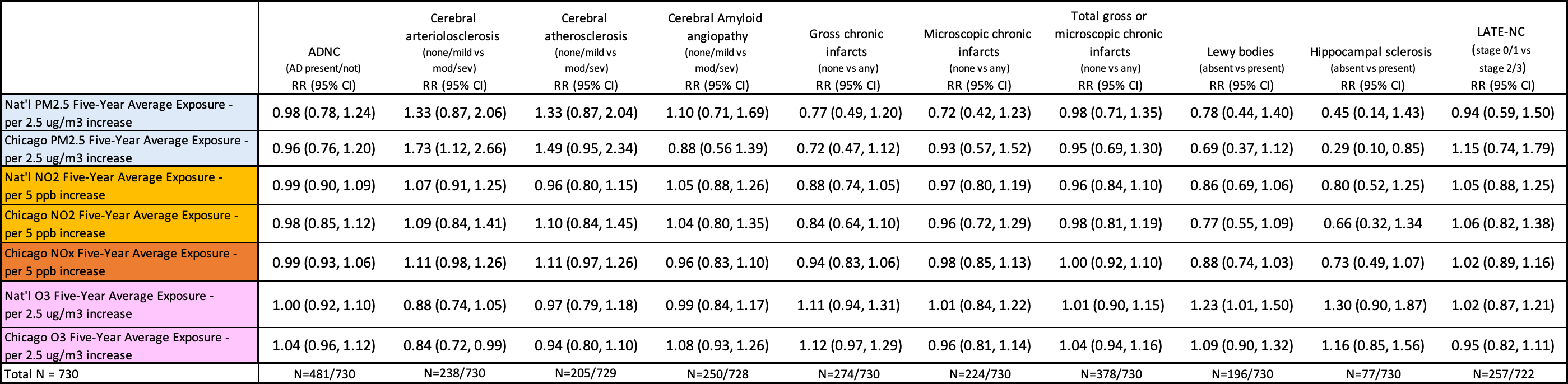

Exposures computed over the 5 years prior to death, using national and Chicago prediction models. N = events/total sample size for dichotomous outcomes. We used log binomial regression models to estimate prevalence ratios per exposure increment of ADNC, cerebral arteriolosclerosis, cerebral atherosclerosis, cerebral amyloid angiopathy, vascular chronic infarcts (gross, microscopic, and total), Lewy bodies, hippocampal sclerosis, and LATE-NC. Models were adjusted for time to autopsy, birth year, death year, sex, race/ethnicity, years of education, baseline smoking status, baseline income, income at age 40, early life socioeconomic status, ≥ 1 ApoE4 allele. ADNC: Alzheimer’s disease neuropathology. RADC: Rush Alzheimer’s Disease Center. MAP: Memory and Aging Project. MARS: Minority Aging Research Study. LATC: Latino Core, RCC: Rush Clinical Core. LATE-NC: limbic predominant age-related TDP-43 encephalopathy - neuropathologic change.

**Table S9.** Results from sensitivity analysis (b). Estimated mean difference and 95% confidence intervals (CI) in beta-amyloid density and tau tangle density at autopsy per increment in 5-year average exposure to national- and Chicago-modeled air pollutants, in which we used exposures estimated over the first three years of the five-year window prior to death. RADC Cohorts: MAP, MARS, LATC, RCC.

|  | Mean difference (95% CI) | | |
| --- | --- | --- | --- |
|  | Beta-amyloid density | Tau tangle  density | |
| Nat'l PM2.5 Five-Year Average  Exposure - per 2.5 ug/m3 increase* | -0.04 (-0.26, 0.18) | | 0.16 (-0.11, 0.44) |
| Chicago PM2.5 Five-Year Average Exposure - per 2.5 ug/m3 increase** | -0.16 (-0.39, 0.06) | | 0.19 (-0.09, 0.47) |
| Nat'l NO2 Five-Year Average**  Exposure - per 5 ppb increase | -0.02 (-0.11, 0.07) | | 0.05 (-0.06, 0.16) |
| Chicago NO2 Five-Year Average** Exposure - per 5 ppb increase | -0.15 (-0.29, -0.001) | | 0.15 (-0.03, 0.34) |
| Chicago NOx Five-Year Average** Exposure - per 5 ppb increase | -0.10 (-0.17, -0.04) | | 0.04 (-0.04, 0.13) |
| Nat'l O3 Five-Year Average***  Exposure - per 2.5 ppb increase | 0.00 (-0.08, 0.09) | | -0.04 (-0.15, 0.07) |
| Chicago O3 Five-Year Average**** Exposure - per 2.5 ppb increase | 0.06 (-0.03, 0.15) | | 0.01 (-0.12, 0.10) |
| *Total N = 710 (Nat'l PM2.5) | N=709 | | N=707 |
| **Total N = 659 (Chi PM2.5/NO2/NOx) | N=658 | | N=656 |
| *** N = 764 (Nat'l NO2/O3) | N=763 | | N=761 |
| ****N = 637 Chi O3 | N=636 | | N=634 |

Exposures computed for the three years prior to a two-year lag prior to death, using national and Chicago prediction models. N = total sample size for. Beta-amyloid (Aβ) density was measured as the square root-transformed mean percent of Aβ positivity in the cortex. Tau tangle density was measured as square root-transformed mean of tau-tangles per square mm. We used linear regression models to estimate the mean difference per exposure increment in β-amyloid and tau tangle densities. Models were adjusted for time to autopsy, birth year, death year, sex, race/ethnicity, years of education, baseline smoking status, baseline income, income at age 40, early life socioeconomic status, ≥ 1 ApoE4 allele. RADC: Rush Alzheimer’s Disease Center. MAP: Memory and Aging Project. MARS: Minority Aging Research Study. LATC: Latino Core, RCC: Rush Clinical Core.

**Table S10**. Risk ratios (RR) and 95% confidence intervals (CI) for association of 5-year average exposure to national- and Chicago-modeled air pollutants with ADNC, cerebral arteriolosclerosis, cerebral atherosclerosis, cerebral amyloid angiography, chronic cerebrovascular infarctions, Lewy bodies, hippocampal sclerosis, and LATE-NC at autopsy, in which we used exposures estimated over the first three years of the five-year window prior to death. RADC Cohorts: MAP, MARS, LATC, RCC.

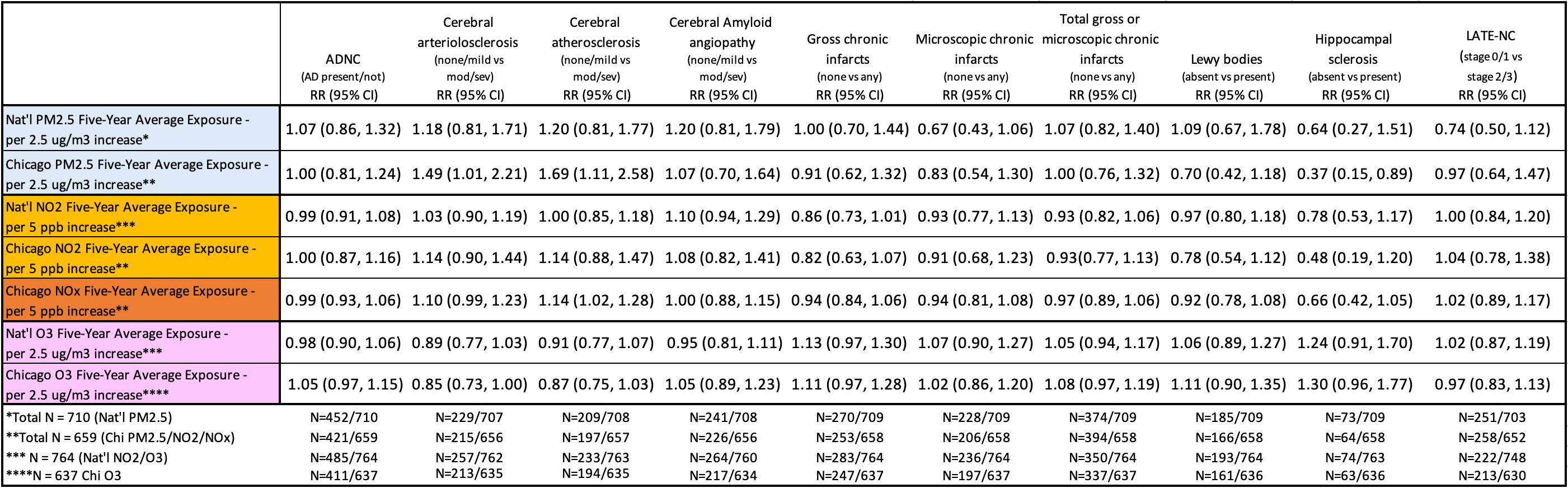

Exposures computed for the three years prior to a two-year lag prior to death, using national and Chicago prediction models. N = events/total sample size for dichotomous outcomes. We used log binomial regression models to estimate prevalence ratios per exposure increment of ADNC, cerebral arteriolosclerosis, cerebral atherosclerosis, cerebral amyloid angiopathy, vascular chronic infarcts (gross, microscopic, and total), Lewy bodies, hippocampal sclerosis, and LATE-NC. Models were adjusted for time to autopsy, birth year, death year, sex, race/ethnicity, years of education, baseline smoking status, baseline income, income at age 40, early life socioeconomic status, ≥ 1 ApoE4 allele. ADNC: Alzheimer’s disease neuropathology. RADC: Rush Alzheimer’s Disease Center. MAP: Memory and Aging Project. MARS: Minority Aging Research Study. LATC: Latino Core. RCC: Rush Clinical Core.

1. All participants who underwent autopsy were included in analyses of NO_2.5_ and O_3_, estimated by the national air pollution models, in association with ADNC. For analyses of other exposure-outcome pairs, the missingness of exposure and/or outcome data was largely due to temporal misalignment of date of death with an exposure model and/or to changes over time in measures included in the autopsy protocol.
